# Low dose mRNA-1273 COVID-19 vaccine generates durable T cell memory and antibodies enhanced by pre-existing crossreactive T cell memory

**DOI:** 10.1101/2021.06.30.21259787

**Authors:** Jose Mateus, Jennifer M. Dan, Zeli Zhang, Carolyn Rydyznski Moderbacher, Marshall Lammers, Benjamin Goodwin, Alessandro Sette, Shane Crotty, Daniela Weiskopf

**Affiliations:** Center for Infectious Disease and Vaccine Research, La Jolla Institute for Immunology (LJI), La Jolla, CA 92037, USA; Department of Medicine, Division of Infectious Diseases and Global Public Health, University of California, San Diego (UCSD), La Jolla, CA 92037, USA

## Abstract

Understanding human immune responses to SARS-CoV-2 RNA vaccines is of interest for a panoply of reasons. Here we examined vaccine-specific CD4^+^ T cell, CD8^+^ T cell, binding antibody, and neutralizing antibody responses to the 25 μg Moderna mRNA-1273 vaccine over 7 months post-immunization, including multiple age groups, with a particular interest in assessing whether pre-existing crossreactive T cell memory impacts vaccine-generated immunity. Low dose (25 μg) mRNA-1273 elicited durable Spike binding antibodies comparable to that of convalescent COVID-19 cases. Vaccine-generated Spike memory CD4^+^ T cells 6 months post-boost were comparable in quantity and quality to COVID-19 cases, including the presence of T_FH_ cells and IFNγ-expressing cells. Spike CD8^+^ T cells were generated in 88% of subjects, with equivalent percentages of CD8^+^ T cell memory responders at 6 months post-boost compared to COVID-19 cases. Lastly, subjects with pre-existing crossreactive CD4^+^ T cell memory had increased CD4^+^ T cell and antibody responses to the vaccine, demonstrating a biological relevance of SARS-CoV-2 crossreactive CD4^+^ T cells.

**One-Sentence Summary:** The mRNA-1273 vaccine induces a durable and functional T cell and antibody response comparable to natural infection.

## Main Text

mRNA vaccines have demonstrated impressive protection from COVID-19 (*1-7*). The COVID-19 vaccine mRNA-1273 encodes a stabilized SARS-CoV-2 full length Spike, developed as a collaboration between Moderna and the NIH Vaccine Research Center (*8, 9*). Durability of immunity has been, and remains, a major unknown for mRNA vaccines in humans. Encouraging reports from both Pfizer/BioNTech and Moderna indicate protective immunity of 90-91% for 6 months after the 2^nd^ immunization (seven months after 1^st^ immunization) (*10, 11*) only down slightly from the 95% maximal protection observed for each of those two vaccines within 1-2 months after two immunizations (*1, 2*). While neutralizing antibodies are a clear correlate of immunity after two immunizations (*12*), the mechanisms of immunity remain unclear, and it remains unknown whether those mechanisms of immunity change as the immune response develops (e.g., after a single immunization) or as immune memory changes composition (*12-14*). Direct measurements of immune memory compartments in humans are necessary to provide insights into these important topics.

Infection and vaccination are two different paths to immunity. Comparison of vaccine-generated immune memory to immune memory of persons infected with SARS-CoV-2 is of value, as studies have indicated natural immunity is 93-100% protective against symptomatic reinfection for 7-8 months (*15-18*); though natural immunity protection against certain variants of concern (VOCs) is likely to be lower (*19*). After SARS-CoV-2 infection, immunological memory has been observed for 8+ months for CD4^+^ T cells, CD8^+^ T cells, memory B cells, and antibodies (*20, 21*). The immune memory in response to SARS-CoV-2 infection exhibits a relatively gradual decline that partially stabilizes within 1 year (*22-25*). It is unknown whether immune memory to mRNA-1273 vaccine months after immunization is similar or different to memory generated by SARS-CoV-2 infection.

Pre-existing crossreactive memory CD4^+^ T cells that recognize SARS-CoV-2 have been found in ∼50% of individuals, pre-pandemic (*26-33*). There has been intense interest in understanding whether these pre-existing crossreactive memory CD4^+^ T cells, identified in vitro, are biologically relevant in v*ivo (34, 35)*. One approach to test the relevance of such T cells in a controlled fashion is in the context of a vaccine trial, as individuals in a clinical trial are all exposed to a well-defined dose of antigen at a specific time, such that the presence of pre-existing crossreactive T cell memory can be measured immediately prior to the new antigen exposure. Additionally, a low dose of antigen exposure may be more sensitive for detection of an impact of crossreactive memory on immune responses.

An open-label, age de-escalation Phase 1 trial utilized the mRNA-1273 vaccine with immunizations on day 1 and 29 (*9, 36*). To characterize the antibody response over 7 months elicited by two 25 µg doses of mRNA-1273 COVID-19 vaccine, we quantified SARS-CoV-2 Spike and receptor binding domain (RBD) immunoglobulin G (IgG). In addition, SARS-CoV-2 pseudovirus (PSV) neutralization titers were determined. All analyses were performed with samples collected on study day 1, 15, 43, and 209. Spike IgG titers were significantly higher at day 15, 43, and 209 compared to day 1 (*p*<0.0001, **Fig. 1A**). Spike IgG was maintained at detectable levels for at least 7 months from the 1^st^ vaccination, for 100% (33/33) of subjects. Likewise, RBD IgG titers were significantly higher at all three timepoints following vaccination (*p*<0.0001, **Fig. 1B**). RBD IgG was induced by one immunization in 94% (33/35) of subjects. This response rate increased to 100% (33/33) after the 2^nd^ immunization, and was maintained for at least 6 months after the 2^nd^ vaccination, consistent with the Spike IgG. SARS-CoV-2 PSV neutralizing titers were significantly higher at all three post-vaccination timepoints (*p<*0.001, **Fig. 1C**). PSV neutralizing titers were detected in 29% (10/35) of subjects after one vaccination, 100% after two vaccinations (33/33), and 88% (29/33) subsequently maintained detectable neutralizing antibodies for at least 6 months after the 2^nd^ vaccination.

**Fig. 1.**
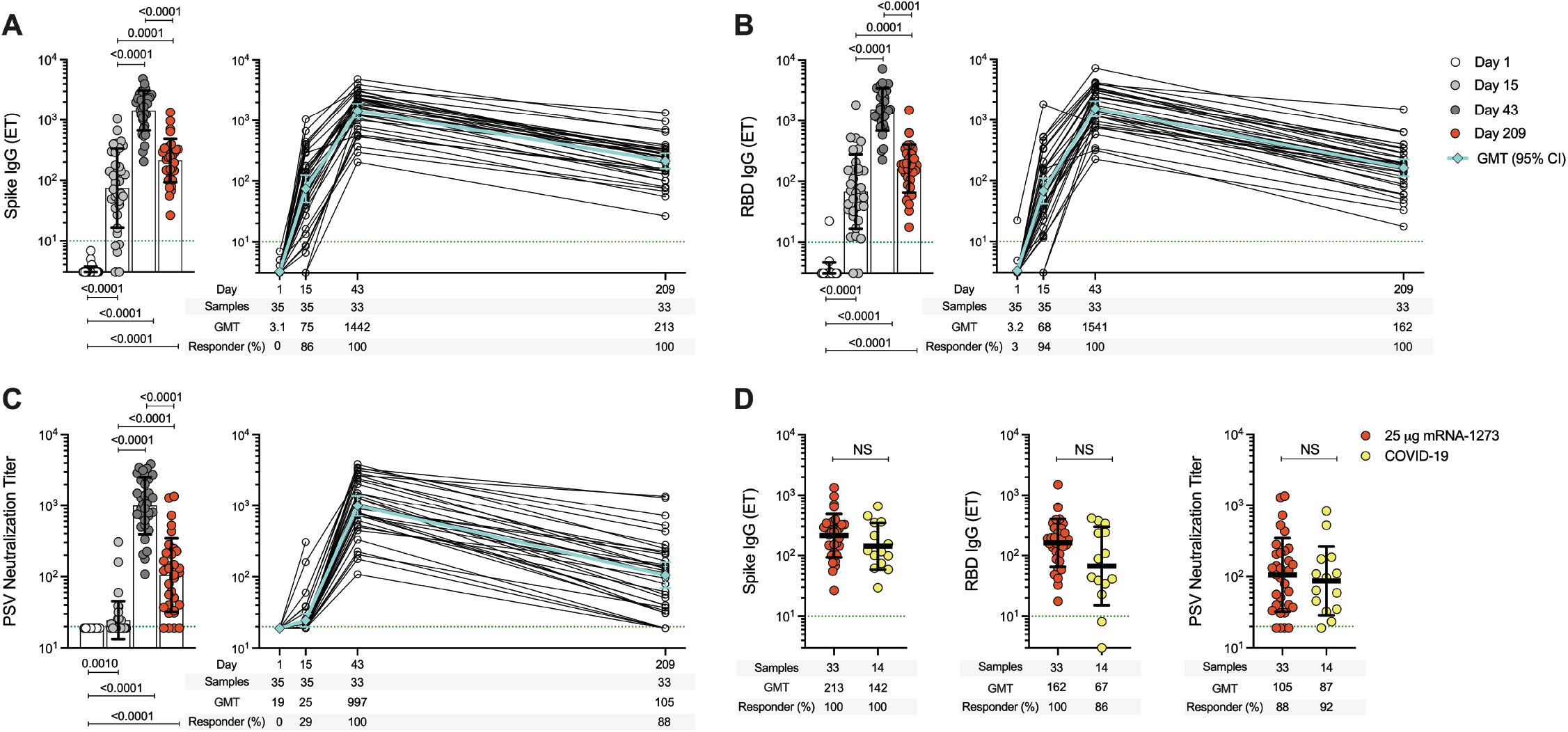
Circulating SARS-CoV-2 Spike specific antibodies induced by 25 μg mRNA-1273 vaccination. **(A)** Longitudinal SARS-CoV-2 Spike IgG binding titers **(B)** Longitudinal SARS-CoV-2 RBD IgG binding titers and **(C)** Longitudinal SARS-CoV-2 Spike Pseudovirus neutralizing titers (PSV). Participants received two injections of the 25 μg mRNA-1273 vaccine, 28 days apart. PBMC samples were collected on day 1, day 15 ± 2 (two weeks post 1^st^ dose), day 43 ± 2 (two weeks post 2^nd^ dose), and day 209 ± 7 days (6 months post 2^nd^ dose). **(D)** Comparison of Spike IgG, RBD IgG, and PSV neutralizing titers induced by two doses of 25 μg mRNA-1273 vaccine at day 209 ± 7 (red circles, n=33) and COVID-19 convalescent donors at 170-195 days PSO (yellow circles, n=14). The dotted green line indicates limit of quantification (LOQ). The bars in A, B, and C indicate the geometric mean titers (GMT) for Spike IgG (Endpoint titers ET), RBD IgG (ET), and PSV neutralizing titers, respectively, on day 1, 15 ± 2, 43 ± 2, and 209 ± 7 postimmunization. Background-subtracted and log data analyzed in all cases. The bottom panels in A-D show the samples included in each group, the GMT, and the percentage of responders. Day 1 = white, day 15 ± 2 = light gray, day 43 ± 2 = dark gray, day 209 ± 7 = red. Statistics by (A, B, and C) Wilcoxon signed-rank test and (D) Mann-Whitney U test. NS, non-significant.

All three measures of antibody response demonstrated similar kinetics (**Fig. 1A-C**) and were highly correlated (r=0.89-0.90, **Fig. S1**). Spike IgG, RBD IgG, and PSV titers at 7 months (study day 209; 181 days after the 2^nd^ immunization) were 6.8-fold, 9.5-fold, and 9.5-fold lower than peak titers. The 25 μg mRNA-1273 vaccine generated antibodies were compared to antibodies from subjects with previous COVID-19 cases collected at 7 months post-symptom onset (PSO, 170-195 days), with no significant differences detected (**Fig. 1D**). Overall, the serological data demonstrated that significant Spike IgG, RBD IgG and neutralizing antibody responses were induced in response to two 25 µg doses of mRNA-1273 and maintained in 88-100% of vaccinees for at least 6 months after the 2^nd^ immunization, and resemble the levels observed after natural infection with SARS-CoV-2.

Spike-specific CD4^+^ T cell responses were measured utilizing a flow cytometry activation induced marker (AIM) assay (*37*) and a peptide pool containing overlapping 15-mers encompassing the entire SARS-CoV-2 Spike sequence (*27, 38*). Antigen-specific CD4^+^ T cells were identified based on double positivity for CD137^+^ and OX40^+^ (**Fig. S2**). Spike-specific CD4^+^ T cell frequencies were significantly higher at day 15, 43, and 209 compared to day 1 (*p*<0.0001, **Fig. 2A**). On day 1, before vaccination, Spike-specific CD4^+^ T cells with a predominantly memory phenotype were detected in 49% of clinical trial subjects (17/35), demonstrating the presence of pre-existing SARS-CoV-2 Spike crossreactive memory CD4^+^ T cells (*26-30, 33*), as discussed in the latter part of this report. Spike-specific CD4^+^ T cell responses were observed after the 1^st^ vaccination in 97% of subjects (34/35, **Fig. 2A**). CMV-specific CD4^+^ T cells were unchanged, as expected, indicating no bystander influence of the mRNA-1273 vaccination (**Fig. S3**). The SARS-CoV-2 Spike CD4^+^ T cell response rate increased to 100% (32/32) after the 2^nd^ vaccination and was then maintained for at least 6 months after the 2^nd^ vaccination. Spike-specific memory CD4^+^ T cell frequencies 7 months after the 1^st^ vaccination were similar to that observed for COVID-19 cases (COVID-19 samples collected 170-195 days PSO, **Fig. 2B**). Median mRNA-1273 vaccine-generated Spike-specific CD4^+^ T cell frequencies at all time points post-vaccination also exceeded those of CMV-specific CD4^+^ T cells (**Fig. 2A, Fig. S3A**).

**Fig. 2.**
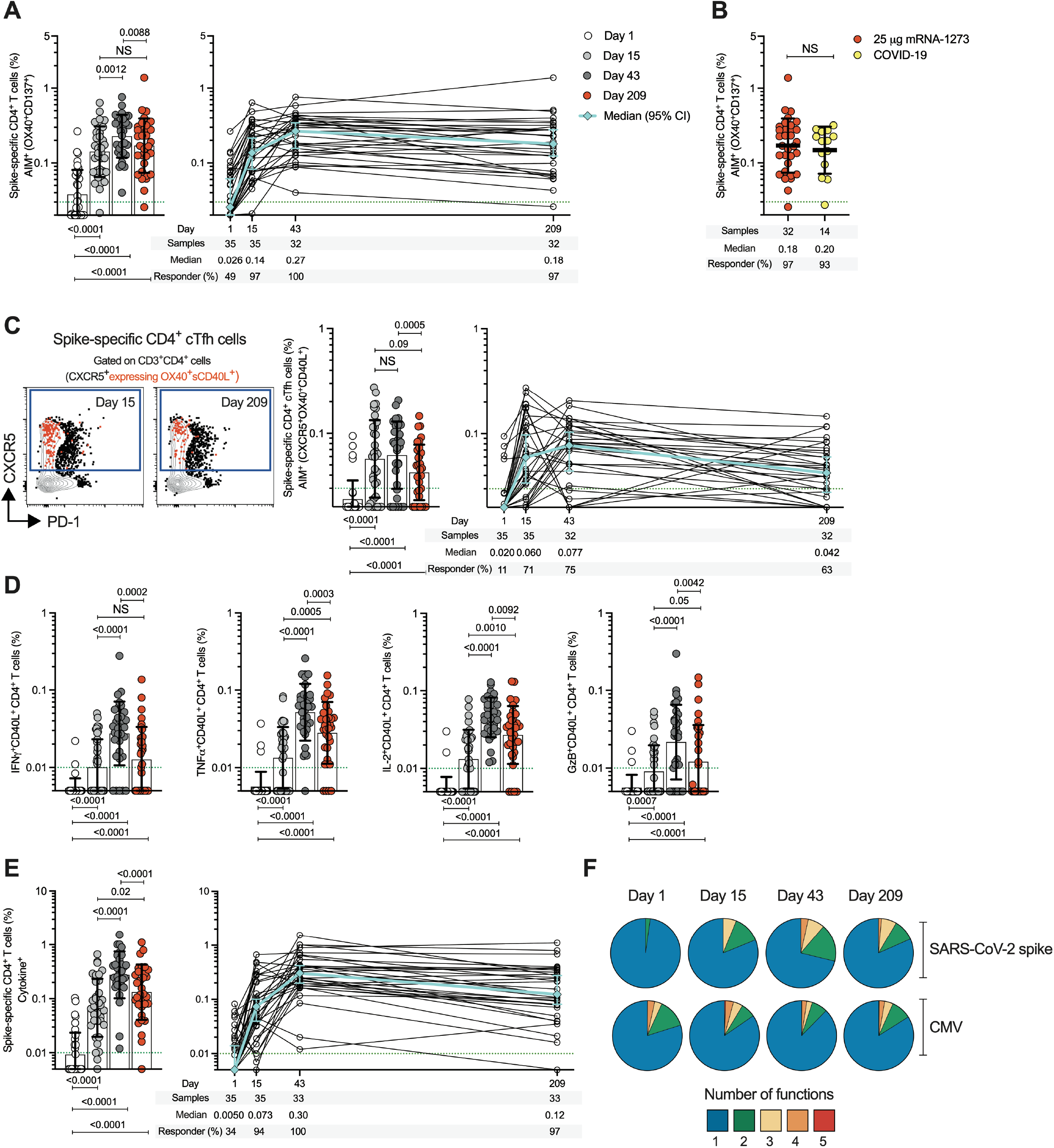
mRNA-1273 vaccination induces durable and multifunctional Spike-specific CD4^+^ T cell responses. **(A)** Longitudinal Spike-specific CD4^+^ AIM^+^ T cells in mRNA-1273 vaccinees. Percentage of background subtracted Spike-specific CD4^+^ T cells quantified by AIM (OX40^+^ CD137^+^) after 24 h stimulation with overlapping Spike megapool (MP) in mRNA-1273 vaccinees (See **Fig. S2** for gating strategy). **(B)** Comparison of Spike-specific AIM^+^ CD4^+^ T cell frequencies between 25 μg mRNA-1273 vaccine at day 209 ± 7 (red circles, n=32) and COVID-19 convalescent donors at 170-195 days PSO (yellow circles, n=14). **(C)** Quantitation of Spike-specific circulating T follicular helper (cT_FH_) cells (CXCR5^+^ OX40^+^ surface CD40L^+^, as % of CD4^+^ T cells) after overnight stimulation with Spike MP. Representative gating strategy of CD4^+^ T cells expressing CXCR5^+^ OX40^+^ sCD40L+ from a single vaccinee at day 15 ± 2 and day 209 ± 7. **(D)** Spike-specific CD4^+^ T cells expressing intracellular CD40L (iCD40L) and producing IFNγ, TNFα, IL-2 or GzB in mRNA-1273 vaccinees. **(E)** Longitudinal Spike-specific CD4^+^ Cytokine^+^ T cells expressing iCD40L and/or IFNγ, TNFα, IL-2 or GzB in mRNA-1273 vaccinees. Percentage of background subtracted Spike-specific CD4^+^ T cells quantified by ICS after stimulation with Spike MP in mRNA-1273 vaccinees (See **Fig. S4** for gating strategy). A Boolean gating strategy was used to define the frequencies of CD4^+^ T cells producing either IFNγ, TNFα, IL-2 or GzB or expressing iCD40L. The dotted green line indicates limit of quantification (LOQ). The bars in A, C, D, and E indicate the geometric mean in the analysis of the Spike-specific CD4^+^ T cells on day 1, 15 ± 2, 43 ± 2, and 209 ± 7 postimmunization. Background-subtracted and log data analyzed in all cases. The bottom panels in A, B, C, and E show the samples included in each group, the median of the frequencies, and the percentage of responders. Day 1 = white, day 15 ± 2 = light gray, day 43 ± 2 = dark gray, day 209 ± 7 = red. **(F)** Longitudinal Multifunctional Spike-specific CD4^+^ T cells in mRNA-1273 vaccinees. Proportions of multifunctional activity profiles of the Spike– specific CD4^+^ T cells from mRNA-1273 vaccinees evaluated at day 1, day 15 ± 2, day 43 ± 2, and day 209 ± 7. The blue, green, yellow, orange, and red colors in the pie charts depict the production of one, two, three, four, and five functions, respectively. Statistics by (A, C, D, and E) Wilcoxon signed-rank test and (B) Mann-Whitney U test. NS, non-significant.

To assess functionality, we measured T follicular helper (T_FH_) differentiation and cytokine production by the vaccine-generated Spike-specific CD4^+^ T cells. T_FH_ cells are the specialized subset of CD4^+^ T cells required for B cell help and are critical for the generation of neutralizing antibodies in most conditions (*39*). Spike-specific circulating T_FH_ cell (cT_FH_) frequencies were significantly higher at days 15, 43, and 209 compared to day 1 (*p*<0.0001, **Fig. 2C**, right panel). Spike-specific cT_FH_ were detected in 71% (25/35) and 75% (24/32) of subjects after the 1^st^ and 2^nd^ vaccination, respectively. Spike-specific cT_FH_ responses were detectable in 94% of subjects overall (32/35); different response kinetics were observed at the level of individual donors (**Fig. 2C**, right panel). Spike specific memory cT_FH_ cells were still detected in 63% of vaccinees 6 months after the 2^nd^ vaccination (20/32, **Fig. 2C**).

Vaccine-specific CD4^+^ T cell cytokine profiles were determined by flow cytometry based intracellular IFNγ, TNFα, IL-2 or granzyme B (GzB) expression among intracellular CD40L^+^ (iCD40L) cells after stimulation with SARS-CoV-2 Spike peptides (**Fig. S4**). Cytokine-producing Spike-specific CD4^+^ T cell frequencies were significantly higher at days 15, 43, and 209 compared to day 1 (*p*<0.0001, **Fig. 2D**). After the 2^nd^ vaccination (day 43), Spike-specific CD4^+^ IFNγ^+^ cells were detected in 89% (28/33), TNFα^+^ cells in 97% (32/33), IL-2^+^ cells in 100% (33/33), and GzB^+^ cells in 76% (25/33) of subjects, respectively (**Fig. 2D**). No to little IL-4, IL-17A, or IL-10 was detected at any timepoint (**Fig. S5)**. Next, we applied a Boolean gating strategy to define the multifunctionality of Spike-specific CD4^+^ T cells, quantifying CD40L^+^ cells producing IFNγ, TNFα, IL-2, and/or GzB (**Fig. S6, Fig. 2E**). Cytokine-producing Spike-specific CD4^+^ T cells were observed in 94% (33/35) and 100% (33/33) of subjects after the 1^st^ and 2^nd^ vaccination, respectively, and were maintained for at least 6 months after the 2^nd^ vaccination (97%, 32/33 of the subjects. **Fig. 2E**). Multifunctional CD4^+^ T cells were observed (**Fig. 2F**), with similar proportions of multifunctionality seen for vaccine-generated Spike-specific cells compared to CMV-specific cells (**Fig. 2F, Fig. S3C**). Spike-specific multifunctional memory CD4^+^ T cells were maintained for at least 6 months after the 2^nd^ vaccination (**Fig. 2F**). Overall, these data demonstrate that robust Spike-specific CD4^+^ T cells were generated by low dose mRNA-1723 vaccine. The memory CD4^+^ T cell functionality profile indicates T_FH_ and TH1 polarization (**Fig. 2C-F**), with an absence of TH2 cells and a presence of multifunctional cells, which is a profile desirable for antiviral immunity.

SARS-CoV-2 Spike-specific CD8^+^ T cell responses were measured by AIM (CD69^+^ and CD137^+^, **Fig. S2**) using the pool of overlapping 15-mers spanning the entire Spike sequence. Spike-specific CD8^+^ T cell frequencies were significantly higher at all three timepoints post-vaccination (*p*<0.0005, **Fig. 2A**), with responses observed in 34% (12/35) and 53% (17/32) of subjects after the 1^st^ and 2^nd^ vaccination, respectively (**Fig. 3A**). Spike-specific CD8^+^ T cells were detectable for at least 6 months after the 2^nd^ vaccination, with a response rate comparable to that observed for COVID-19 cases (**Fig. 3B**). The CD8^+^ T cell cytokine production was measured (IFNγ, TNFα, IL-2, or GzB, **Fig. S8**); IFNγ and TNFα expression was significantly higher at study day 15, 43, and 209 compared to day 1 (*p*<0.001, **Fig. 2C**). At day 43, Spike-specific CD8^+^ cytokine responses were detected in 70% (23/33) for IFNγ, 39% (13/33) for TNFα, and 12% (4/33) for IL-2. Multiple positive and negative controls samples and experimental conditions were used to demonstrate the specificity of the CD8^+^ T cell responses (**Fig. 3, Fig. S2, Fig. S4E-G**). We compared Spike-specific T cell responses detected by the AIM and ICS methods on days 15, 43, and 209. Correlation between the two methods was highly significant for both Spike-specific CD4^+^ and CD8^+^ T cells (*p*<0.0001; **Fig. S7**). A Boolean gating strategy was also used to assess multifunctionality of cytokine-secreting Spike-specific CD8^+^ T cells (**Fig. S8, Fig. 3D**). The 1^st^ immunization induced significant Spike-specific CD8^+^ cytokine responses in 51% (18/35) of subjects, increasing to 70% (23/33) of subjects after the 2^nd^ vaccination **(Fig. 3D**). The fraction of multifunctional Spike-specific CD8^+^ T cells also increased between 1^st^ and 2^nd^ vaccination. The most prevalent profile of CD8^+^ T cells with three functions was GzB^+^IFNγ^+^TNFα^+^ (**Fig. S8**), similar to that of CMV-specific CD8^+^ T cells (**Fig. 3D, S9**). Overall, the data demonstrate that 25 µg mRNA-1273 vaccination induced multifunctional Spike-specific memory CD8^+^ T cells.

**Fig. 3.**
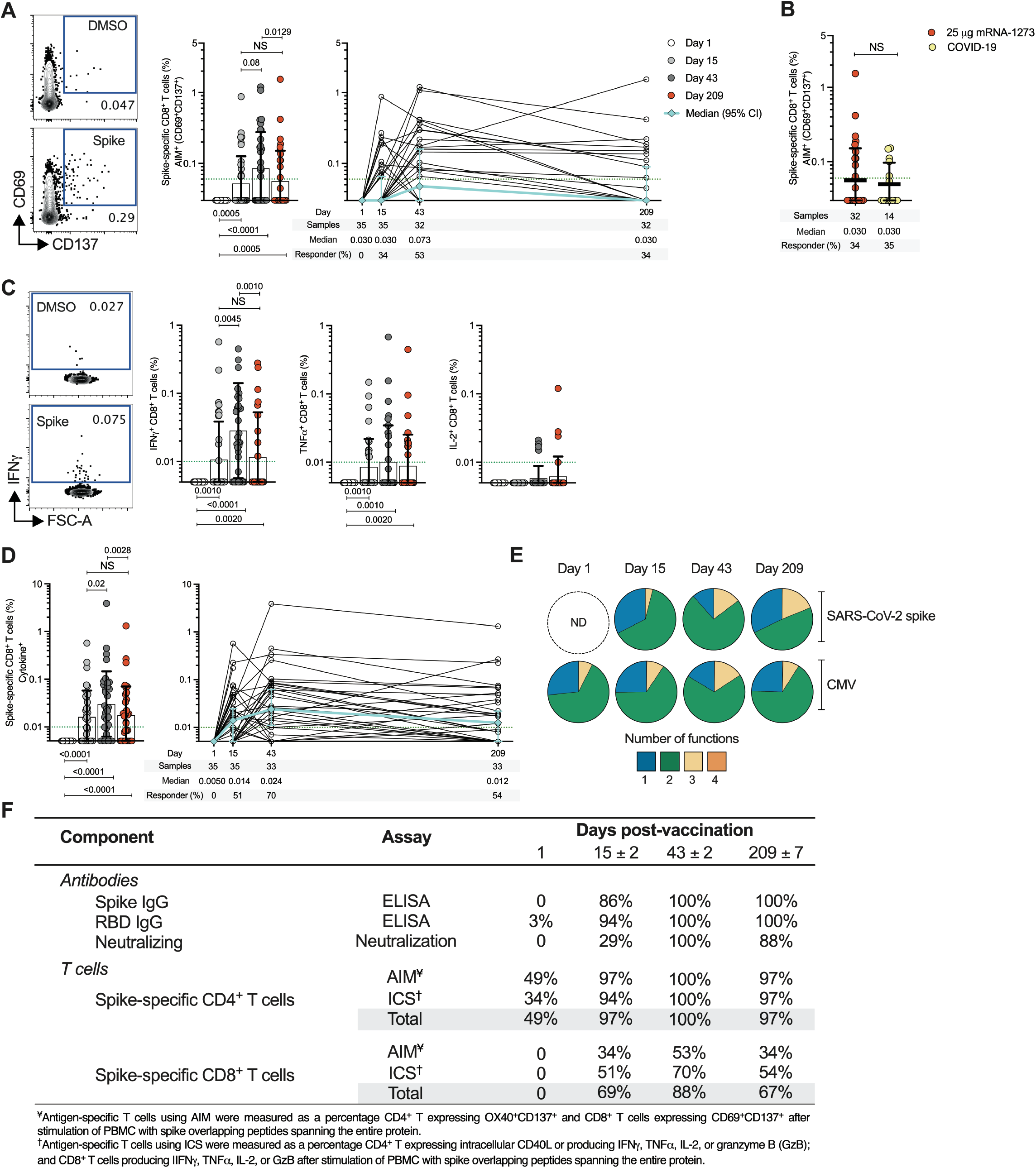
mRNA-1273 vaccination induces multifunctional Spike-specific CD8^+^ T cell responses. **(A)** Longitudinal Spike-specific CD8^+^ AIM^+^ T cells in mRNA-1273 vaccinees. Representative examples of flow cytometry plots of Spike-specific CD8^+^ T cells (CD69^+^ CD137^+^, after overnight stimulation with Spike MP), compared to DMSO (Left panel). Percentage of background subtracted Spike-specific CD8^+^ T cells quantified by AIM (CD69^+^ CD137^+^) in mRNA-1273 vaccinees (Right panel, see **Fig. S2** for gating strategy). **(B)** Comparison of Spike-specific AIM^+^ CD8^+^ T cell frequencies between 25 μg mRNA-1273 vaccinees at day 209 ± 7 (red circles, n=32) and COVID-19 convalescent donors at 170-195 days PSO (yellow circles, n=14). Spike-specific CD8^+^ T cells producing IFNγ, TNFα, and IL-2 in mRNA-1273 vaccinees. **(D)** Longitudinal Spike-specific CD8^+^ Cytokine^+^ T cells producing IFNγ, TNFα, IL-2, and GzB in mRNA-1273 vaccinees. Percentage of background subtracted Spike-specific CD8^+^ T cells quantified by Intracellular cytokine staining (ICS) after stimulation with Spike MP in mRNA-1273 vaccinees (See **Fig. S4** for gating strategy). A Boolean gating strategy was used to define the frequencies of CD8^+^ T cells producing either IFNγ, TNFα, IL-2 or GzB. The dotted green line indicates limit of quantification (LOQ). The bars in A, C, and D indicate the geometric mean in the analysis of the Spike-specific CD8^+^ T cells on day 1, 15 ± 2, 43 ± 2, and 209 ± 7 postimmunization. Background-subtracted and log data analyzed in all cases. The bottom panels in A, B, and D show the samples included in each group, the median of the frequencies, and the percentage of responders. Day 1 = white, day 15 ± 2 = light gray, day 43 ± 2 = dark gray, day 209 ± 7 = red. **(E)** Multifunctional Spike-specific CD8^+^ T cells in mRNA-1273 vaccinees. Proportions of multifunctional activity profiles of the Spike–specific CD8^+^ T cells from mRNA-1273 vaccinees evaluated at day 1, day 15 ± 2, day 43 ± 2, and day 209 ± 7. The blue, green, yellow, and orange colors in the pie charts depict the production of one, two, three, and four functions, respectively. **(F)** Spike-specific immune responses detected in 25 μg mRNA-1273 vaccinees. The table shows the percentage of responder for antibody and T cell responses. The overall Spike-specific T cell response was calculated based on the AIM and ICS results per donor and time-point. Statistics by (A, C, and D) Wilcoxon signed-rank test and (B) Mann-Whitney U test. NS, non-significant, ND, non-detectable, ELISA, Enzyme-linked immunosorbent assay; AIM, Activation induced markers; ICS, Intracellular cytokine staining.

The vaccine immune response data, summarized in **Figure 3F**, demonstrate that Spike antibody, CD4^+^ and CD8^+^ T cell responses generated by 25 µg mRNA-1273 vaccination are multifunctional, durable, and comparable in magnitude to those induced by natural infection. A concern has been raised that vaccination may not induce adequate immune memory in the elderly (*40*). This vaccination cohort consisted of volunteers from three different age groups (*9, 36*). Accordingly, we repeated our analyses dividing the donors based on age 18-55, 56-70, and >70 years of age. While the study size is underpowered for in-depth examination of the three age groups, we found that levels of Spike IgG and RBD antibodies in the older groups were reduced approximately two-fold on day 209 post-vaccination (**Fig. S10A-B**), similar to that recently reported for the 100 μg mRNA-1273 vaccination (*36*). Spike-specific CD4^+^ or CD8^+^ T cells were not reduced in the two older vaccinee groups (56-70 and >70 years) compared to the 18–55-year-old age group. Memory CD4^+^ and CD8^+^ T cell frequencies at 6 months after the 2^nd^ vaccination were at least as strong in the older age groups as the younger adults (**Fig. S10D-H**). In summary, a slight reduction in antibody but not T cell memory was observed in older adults.

Pre-existing crossreactive memory T cells recognizing SARS-CoV-2 in vitro are found in many individuals (*26-33*). It was hypothesized that the existence of pre-existing Spike immune memory might modulate immune responses to infection or vaccination (*32*). To address this question, we separated our cohort as a function of whether each subject possessed pre-existing crossreactive memory CD4^+^ T cells reactive against SARS-CoV-2 Spike (**Fig. 4A-B**). As noted above (**Fig 2A**), we detected pre-existing SARS-CoV-2 Spike-specific CD4^+^ T cells in 49% (17/35) of the 25 μg mRNA-1273 vaccinees (day1, **Fig. 3F**). After vaccination, Spike-specific CD4^+^ T cells were significantly higher at day 15 in crossreactive memory subjects compared to subjects with no crossreactive memory (2.3-fold; *p*<0.0001, **Fig. 4C**). Spike-specific memory CD4^+^ T cell frequencies remained higher after the 2^nd^ vaccination in subjects with crossreactive memory compared to subjects with no crossreactivity (*p*=0.02, **Fig. 4C**). Six months after the 2^nd^ vaccination, Spike-specific CD4^+^ T cell frequencies remained significantly higher in donors with pre-existing crossreactive memory (*p*=0.01, **Fig. 4C**). Higher frequencies of cytokine-positive Spike-specific CD4^+^ T cells were also observed after the 1^st^ vaccination in the group with crossreactive memory compared to the group without crossreactive memory (*p*=0.0051, **Fig. 4D**). The degree of multifunctionality in the Spike-specific CD4^+^ T cell responses was significantly higher after the first vaccination (*p*=0.02, **Fig. S11, Fig. 4E**). Six months after the 2^nd^ vaccination, no difference in multifunctionality was observed between subjects with and without pre-existing crossreactive immunity (**Fig. S11, Fig. 4D**), suggesting a compensatory effect of booster immunization.

**Fig. 4.**
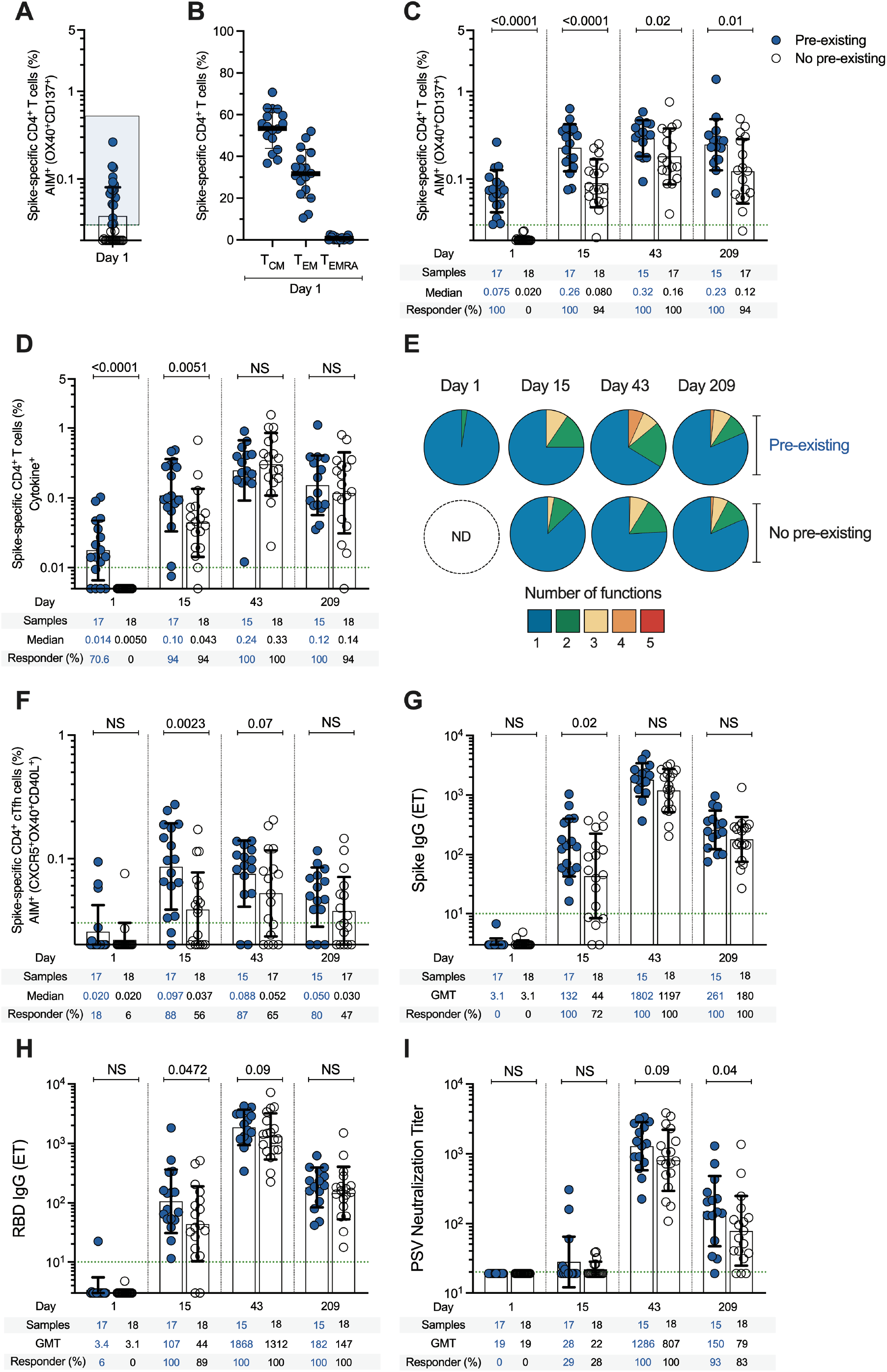
Pre-existing Spike immunity can modulate the T cell and antibody responses after vaccination. **(A)** Pre-existing Spike-specific CD4^+^ AIM^+^ T cells in mRNA-1273 vaccinees at day 1. **(B)** Memory phenotype of pre-existing Spike-specific CD4^+^ AIM^+^ T cells in mRNA-1273 vaccinees at day 1. **(C)** Spike-specific CD4^+^ AIM^+^ T cells in mRNA-1273 vaccinees with and without pre-existing Spike immunity evaluated at day 1, day 15 ± 2, day 43 ± 2, and day 209 ± 7 postimmunization. **(D)** Spike-specific CD4^+^ Cytokine^+^ T cells in mRNA-1273 vaccinees with and without pre-existing Spike immunity evaluated at day 1, day 15 ± 2, day 43 ± 2, and day 209 ± 7 postimmunization. **(E)** Multifunctional Spike-specific CD4^+^ T cells in mRNA-1273 vaccinees with and without pre-existing Spike immunity. Proportions of multifunctional activity profiles of the Spike–specific CD4^+^ T cells from mRNA-1273 vaccinees with and without pre-existing Spike immunity evaluated at day 1, day 15 ± 2, day 43 ± 2, and day 209 ± 7. The blue, green, yellow, orange, and red colors in the pie charts depict the production of one, two, three, four, and five functions, respectively. **(F)** Spike-specific circulating T follicular helper (cT_FH_) cells (CXCR5^+^ OX40^+^ CD40L^+^, as % of CD4^+^ T cells) in mRNA-1273 vaccinees without and with pre-existing Spike immunity at day 1, day 15 ± 2, day 43 ± 2, and day 209 ± 7 postimmunization. **(G)** Spike IgG from mRNA-1273 vaccinees with and without pre-existing Spike immunity. **(H)** RBD IgG from mRNA-1273 vaccinees with and without pre-existing Spike immunity. **(I)** PSV neutralizing titers of SARS-CoV-2 Spike in mRNA-1273 vaccinees with and without pre-existing Spike immunity. The dotted green line indicates limit of quantification (LOQ). The bars in A, C, D, F, G, H, and I indicate the geometric mean in the analysis of the Spike-specific CD4^+^ T cells or antibody titers on day 1, 15 ± 2, 43 ± 2, and 209 ± 7 postimmunization. Background-subtracted and log data analyzed in all cases. The bottom panels in C, D, F, G, H, and I show the samples included in each group, the median of the frequencies or the geometric mean titers (GMT), and the percentage of responders. Vaccinees with pre-existing Spike immunity = blue, vaccinees without pre-existing Spike immunity = clear. Statistics by (C, D, F, G, H, and I) Mann-Whitney U test. NS, non-significant, ND, non-detectable.

The frequencies of T_FH_ cells in subjects with and without crossreactive memory were of particular interest, because of their relevance in antibody responses. 2.6-fold higher frequencies of Spike-specific cT_FH_ cells were observed on day 15 in vaccinees with pre-existing crossreactive Spike-specific CD4^+^ T cell memory (*p*=0.0023, **Fig. 4F**). Likewise, significantly higher levels of Spike IgG (*p*=0.02, **Fig. 4G**) and RBD IgG (*p*=0.047, **Fig. 4H**) were detected on day 15 in vaccinees with pre-existing crossreactive CD4^+^ T cell memory compared to subjects without crossreactive memory. The group with pre-existing crossreactive Spike-specific CD4^+^ T cell memory demonstrated significantly higher neutralizing titers 6 months after the 2^nd^ vaccination, compared to the group without pre-existing crossreactive cells (*p*=0.04, **Fig. 4I**).

Pre-existing crossreactive CD4^+^ T cells were observed in all three age groups (**Fig. S10D)**. We did not detect pre-existing crossreactive Spike-specific CD8^+^ T cell memory (**Fig. 3A, D**; **Fig. S10G**-**H**). We found no modulation of Spike-specific CD8^+^ T cell responses by pre-existing CD4^+^ T cell memory at any timepoint (**Fig. S12**), suggesting that CD4^+^ T cell help to CD8^+^ T cells may not be a primary limiting factor under these vaccination conditions. Crossreactive memory CD4^+^ T cells recognizing epitopes from the non-Spike components of the SARS-CoV-2 proteome were also detected (**Fig. S13**) (*27, 30*). The non-Spike SARS-CoV-2 crossreactive CD4^+^ T cell frequencies were unchanged throughout the four time points investigated, not modulated by mRNA-1273 vaccination (**Fig. S13**), consistent with the vaccine containing only the Spike antigen and not causing bystander activation. Overall, these data demonstrate that pre-existing crossreactive CD4^+^ T cell memory can influence mRNA-1723 vaccine-generated Spike-specific CD4^+^ T cell responses. Higher levels of Spike-specific cT_FH_ responses, as well as Spike specific IgG and RBD-specific IgG levels were detected after a single vaccination in subjects with pre-existing crossreactive memory. Six months after the 2^nd^ vaccination, higher frequencies of vaccine specific memory CD4^+^ T cells and higher titers of neutralizing antibodies were present in individuals with crossreactive CD4^+^ T cell memory.

The SARS-CoV-2 mRNA vaccines have been extraordinary successes. It is important to better understand the immunology of these vaccines in order to (1) better understand mechanisms of protective immunity provided by the vaccines, (2) understand the durability of immune memory (and thus infer trajectories of protective immunity) generated by the vaccines, and (3) understand immunological features of these vaccines that may be relevant for vaccine design against other pathogens. Here, studying 35 vaccinated subjects out at 7 months from the initial immunization, we found that two-dose 25 μg mRNA-1273 vaccination generated immune memory against Spike comparable to that of SARS-CoV-2 infection for antibodies, CD4^+^ T cells, and CD8^+^ T cells. Furthermore, the immune responses were significantly enhanced by the presence of pre-existing crossreactive CD4^+^ T cell memory.

We consistently found Spike-specific memory CD4^+^ T cells in vaccinated subjects 6 months post-2^nd^ low dose mRNA-1273 immunization. Less than a two-fold difference in Spike-specific CD4^+^ T cell frequencies was observed between peak and 6 months post-boost, indicative of a durable memory T cell response to the vaccine. The frequency of memory CD4^+^ T cells was also similar between low dose mRNA-1273 vaccinated persons and COVID-19 cases. The functionality of the mRNA vaccine-elicited memory CD4^+^ T cells was encouraging; the cells exhibited an antiviral functional profile, including substantial representation of T_FH_ cells and IFNγ-expressing cells, and presence of multi-cytokine-expressing cells in proportion similar to CMV-specific memory CD4^+^ T cells.

Uncertainty had surrounded whether the mRNA-1273 COVID-19 vaccine elicited effector CD8^+^ T cells and memory CD8^+^ T cells in humans (*9, 41, 42*). Here we report, at the peak of the immune response, Spike-specific CD8^+^ T cells by AIM or ICS assays were detected in 88% (53% or 70%, respectively) of subjects receiving low dose mRNA-1273, which is a CD8^+^ T cell response rate equivalent to that of COVID-19 cases (**Fig. 3C** and (*20, 21, 27, 43*)). We speculate that absence of detection of Spike-specific CD8^+^ T cells in some studies reflects the stringency of the experimental conditions used. Here, allowance for 24 hours of antigen-stimulation, vaccine-generated CD8^+^ T cells were detected in the majority of individuals. Using peptide:MHC multimers, CD8^+^ T cells were observed to the Pfizer/BioNTech BNT162b2 vaccine at substantial frequencies (*41*). In terms of memory, CD8^+^ T cells were observed two months (64 days) after 2^nd^ immunization with BNT162b2 (*41*). Here, Spike-specific memory CD8^+^ T cells were detected 6 months after the 2^nd^ immunization with 25 μg mRNA-1273. These mRNA-1273 vaccine-generated memory CD8^+^ T cells were detected in 67% of subjects (**Fig. 3F**) and were not dissimilar in magnitude to Spike-specific memory CD8^+^ T cells generated by COVID-19 cases. Thus, both CD4^+^ and CD8^+^ T cell memory is generated by the COVID-19 mRNA-1273 vaccine.

Low dose RNA vaccines have potential advantages for future needs and applications. Dose sparing is one. Low dose immunization is also less reactogenic (*9, 41*). It is of interest to consider different vaccine doses across age groups, or high- versus low-risk groups, but a better understanding of immune memory to different doses is key for such considerations. Data reported here are encouraging demonstrations of the potential of RNA vaccines to generate immune memory, including at lower doses.

Pre-existing immunity in the form of crossreactive memory CD4^+^ T cells had a measurable impact on immune response to the mRNA COVID-19 vaccine in this cohort. This indicates that crossreactive memory T_FH_ cells may both accelerate B cell priming and antibody responses to a new antigen, and also increase the robustness of the long-term humoral immunity, as evidenced by the higher antibody titers. Both total Spike-specific CD4^+^ T cells and Spike-specific T_FH_ cells were enhanced after one immunization in persons with pre-existing crossreactive memory CD4^+^ T cells recognizing SARS-CoV-2 Spike, suggesting that the crossreactive memory CD4^+^ T cells are recalled upon vaccination and impact the CD4^+^ T cell repertoire in response to a Spike vaccine, such as mRNA-1273. The pre-existing immunity findings do not represent the exact scenario of SARS-CoV-2 infection, and therefore it remains unknown whether pre-existing T cells have a biological function during human SARS-CoV-2 infection (*29*). Nevertheless, these data provide evidence that the crossreactive CD4^+^ T cells are biologically relevant in the context of vaccination. Thus, it is plausible that the presence and magnitude of crossreactive memory T cells could accelerate the speed and magnitude of CD4^+^ T cell and antibody responses to SARS-CoV-2 infection, compared to persons who have undetectable levels of crossreactive memory T cells. Early T cell responses have been linked to less severe clinical outcomes (*44, 45*). Limitations of this study include the relatively small sample size and limited cell availability.

## Data Availability

Epitope pools utilized in this paper are available to the scientific community upon request and execution of a material transfer agreement (MTA).

## Acknowledgements

This work used samples from the phase 1 mRNA-1273 study (NCT04283461) (*9, 36*). The mRNA-1273 phase 1 study was sponsored and primarily funded by the National Institute of Allergy and Infectious Diseases (NIAID), National Institutes of Health (NIH), Bethesda, MD; in part with federal funds from the NIAID under grant awards UM1AI148373, to Kaiser Washington; UM1AI148576, UM1AI148684, and NIH P51 OD011132, to Emory University; NIH AID AI149644, and contract award HHSN272201500002C, to Emmes. The study was conducted in collaboration with ModernaTX and funding for the manufacture of mRNA-1273 phase 1 material was provided by the Coalition for Epidemic Preparedness Innovation. We would like to thank the LJI Clinical Core, for healthy donor enrollment and blood sample procurement and Dr. Erica Ollman Saphire for providing the Spike plasmids.

## Funding

This work was funded by the NIH NIAID under awards AI142742 (Cooperative Centers for Human Immunology) (A.S., S.C.) and NIH contract Nr. 75N9301900065 (D.W., A.S.). This work was additionally supported in part by LJI Institutional Funds and the NIAID under K08 award AI135078 (J.M.D.)

## Author contributions

Conceptualization: JM, AS, SC, DW; Methodology: JM, JMD, ZZ, CRM, ML, BG, DW; Formal analysis: JM, JMD, ZZ, AS, SC, DW; Investigation: JM, AS, SC, DW; Funding acquisition: AS, SC, DW; Writing: JM, JMD, ZZ, AS, SC, DW; Supervision: AS, SC, DW.

## Competing interests

A.S. is a consultant for Gritstone, Flow Pharma, Arcturus, Immunoscape, CellCarta, OxfordImmunotech and Avalia. S.C has consulted for Avalia, Roche and GSK.

LJI has filed for patent protection for various aspects of T cell epitope and vaccine design work. All other authors declare no conflict of interest.

## Supplementary Materials

Materials and Methods Figs. S1 to S13

Tables S1 to S5

## Notes

### Clinical Trial

NCT04283461

### Author Declarations

The trial was reviewed and approved by the Advarra institutional review board as previously published. All experiments performed at the La Jolla Institute (LJI) were approved by the institutional review boards (IRB) of the La Jolla Institute (IRB#: VD-214).

