## Supplemental Material for "Low dose mRNA-1273 COVID-19 vaccine generates durable T cell memory and antibodies enhanced by pre-existing crossreactive T cell memory"

**This PDF file includes:**

Materials and Methods
Figs. S1 to S13
Tables S1 to S5

### Materials and Methods

Human subjects

*Samples from the phase I trial of 25 µg mRNA-1273 SARS-CoV-2*

A total of 140 peripheral blood samples were obtained from 35 participants who received the 25 µg dose mRNA-1273 SARS-CoV-2 vaccine in a dose escalation, open-label phase 1 trial (mRN-1273 ClinicalTrials.gov number NCT04283461)(*9*). Participants received two injections of the trial vaccine 28 days apart between March and April of 2020 at one of the two study sites, Kaiser Permanente Washington Health Research Institute in Seattle or at the Emory University School of Medicine in Atlanta. Peripheral blood mononuclear cell (PBMC) samples used in this study were collected at the start of the trial (day 1), 14 days after each vaccination (day 15 ± 2 and 43 ± 2) and 7-months after the first dose (day 209 ± 7).

Participants were grouped into three age groups ranging from 18 to 55 (n=15), 56 to 70 (n=10) and >70 years (n=10) of age. (**Table S1)**. Two participants from the 18-55 group received only one dose of the 25 µg mRNA-1273 doses. The day 40 and day 209 samples (both collected post second dose) were excluded from analysis for these two vaccinees. Both sexes were represented (20:15, M: F) and the average age of the participants was 58 years. Participants identified as white (n=34), Hispanic (n=1) or mixed race (Asian/Hispanic, n=1.) An overview of samples analyzed in this study is provided in **Table S1**.
The trial was reviewed and approved by the Advarra institutional review board as previously published (*9*). All experiments performed at the La Jolla Institute (LJI) were approved by the institutional review boards (IRB) of the La Jolla Institute (IRB#: VD-214). Collection and processing of vaccinee PBMC samples was performed at one of the two study location sites as previously described (*9*).

*Samples from convalescent COVID-19 donors*

To compare levels of immune memory responses induced by 25 µg mRNA-1273 SARS-CoV-2 vaccination to immune memory responses induced by natural infection with SARS-CoV-2, we collected blood from individuals that experienced natural infection with SARS-CoV-2. We matched the 7 months (209 days) postvaccination samples with samples from convalescent donors collected on average 181 days (range 170-195) post symptoms onset (PSO). Assuming an average incubation period of 12 days until symptom onset the timepoint post exposure or vaccination is comparable between the cohorts (*46*). To further match ethnicities between the cohorts, we selected 13 samples from Caucasian donors and one sample from a Hispanic donor. Convalescent donors were California residents, who were either referred to the study by a health care provider or self-referred. In the overall cohort, both sexes were represented (10:4, M: F) and the average age of the donors was 35 years (± 16.53). Details of this COVID -19 convalescent cohort are listed in **Table S2.** Seropositivity against SARS-CoV-2 was confirmed by ELISA, as describe below. At the time of enrollment, all COVID-19 convalescent donors provided informed consent to participate in the present and future studies.

*Peripheral blood mononuclear cells (PBMCs) and plasma isolation*

At the La Jolla Institute, whole blood was collected in heparin coated blood bags and centrifuged for 15 min at 1850 rpm to separate the cellular fraction and plasma. The plasma was then carefully removed from the cell pellet and stored at -20°C. PBMCs were isolated by densitygradient sedimentation using Ficoll-Paque (Lymphoprep, Nycomed Pharma, Oslo, Norway) as previously described (*20, 27*). Isolated PBMCs were cryopreserved in cell recovery media containing 10% Dimethyl sulfoxide (DMSO) (Gibco), supplemented with 10% heat inactivated fetal bovine serum (FBS; Hyclone Laboratories, Logan UT), and stored in liquid nitrogen until used in the assays.

Antibody measurements

*SARS-CoV-2 ELISAs*

SARS-CoV-2 ELISA titers in vaccinated and convalescent samples were determined as previously described (*47*). Briefly, Corning 96-well half-area plates (ThermoFisher, Cat. No.3690) were coated with 1 µg/mL SARS-CoV-2 Spike or RBD protein overnight at 4°C. The next day plates were blocked with 3% milk (Skim Milk Powder ThermoFisher, Cat. No. LP0031, by weight/volume) in Phosphate Buffered Saline (PBS) containing 0.05% Tween-20 (ThermoScientific, Cat. No. J260605-AP) for 2 hours at room temperature. Heat-inactivated plasma (30-60 min at 56 °C) was then added to the plates and incubated for 1.5 hours at room temperature. Plates were washed 5 times with 0.05% PBS/Tween. Secondary antibodies were diluted in 1% milk containing 0.05% Tween-20 in PBS. IgG titers were determined using anti-human IgG peroxidase antibody (Hybridoma Reagent Laboratory HP6123-HRP) was used at 1:1,000 dilution. Endpoint titers (ET) were plotted for each sample, using background subtracted data. Negative and positive controls were used to standardize each assay and normalize across experiments. A positive control standard was created by pooling plasma from 6 convalescent COVID-19 donors to normalize between experiments. The Limit of detection (LOD) was defined as 1:3 for IgG. Limit of quantification (LOQ) for vaccinated individuals was established based on uninfected subjects, using plasma from healthy donors never exposed to or vaccinated against SARS-CoV-2.

*Pseudovirus (PSV) Neutralization Assay*

The PSV neutralization assays of vaccinated and convalescent samples were performed as previously described (*20*). Briefly, $2.5x10^4$ VERO cells (ATCC, Cat. No. CCL-81) were seeded in clear flat-bottom 96-well plates (Thermo Scientific, Cat. No. 165305) to produce a monolayer at the time of infection. Recombinant SARS-CoV-2-S-D614G pseudotyped VSV-ΔG-GFP were generated by transfecting HEK293T cells (ATCC, Cat. No. CRL-321) with plasmid phCMV3-SARS-CoV2-Spike kindly provided by Dr. Erica Saphire and then infecting with VSV-ΔG-GFP. Pre-titrated rVSV-SARS-CoV-2-S-D614G was incubated with serially diluted human heat-inactivated plasma at 37°C for 1-1.5 hours before addition to confluent VERO cell monolayers. Cells were incubated for 16 hours at 37°C in 5% $CO_2$ then fixed in 4% paraformaldehyde in PBS pH 7.4 (Santa Cruz, Cat. No. sc-281692) with 10 µg/ml Hoechst (Thermo Scientific, Cat. No. 62249), and imaged using a CellInsight CX5 imager to quantify the total number of cells and infected GFP expressing cells to determine the percentage of infection. Neutralization titers or inhibition dose 50 ($ID_{50}$) were calculated using the One-Site Fit Log IC50 model in Prism 8.0 (GraphPad). As internal quality control to define the variation interassay, three samples were included across the PSV neutralization assays. Samples that did not reach 50% inhibition at the lowest serum dilution of 1:20 were considered as non-neutralizing and the values were set to 19.

Peptide megapools (MP)

We have previously developed the MP approach to allow simultaneous testing of a large number of epitopes. According to this approach, large numbers of different epitopes are solubilized, pooled, and re-lyophilized to avoid cell toxicity problems associated with high concentrations of DMSO typically encountered when single pre-solubilized epitopes are pooled (*48*). Here, were used for ex vivo stimulation of PBMCs for flow cytometry three MPs to evaluate the antigen-specific T cell response against SARS-CoV-2, as described below. Cytomegalovirus (CMV) MP was used as a control against a ubiquitous pathogen in the experiments.

*SARS-CoV-2 MPs*

To characterize SARS-CoV-2-specific T cell response, we utilized three MPs previously described (*27, 49*). First, we used a Spike MP of 253 overlapping peptides spanning the entire sequence of the Spike protein. As this peptide pool consists of peptides with a length of 15 amino acids, both $CD4^+$ and $CD8^+$ T cells have the capacity to recognize this MP (*50*). In addition, to confirm that the Spike-specific $CD8^+$ T cell response observed in the mRNA-1273 vaccinees is also induced in the absence of the Spike-specific $CD4^+$ T cell response, we performed some experiments with an optimal MP of HLA Class-I epitopes (CD8-S MP). This CD8-S MP consists of 197 9- and 10-mers derived Spike peptides that have previously been described to be recognized by $CD8^+$ T cells in SARS-CoV-2 exposed donors (*51*). Lastly, we used a predicted SARS-CoV-2 specific MP (CD4-R) to evaluate the non-Spike response or the remainder of the SARS-CoV-2 genome of 246 HLA Class-II $CD4^+$ epitopes previously described (*27*). We have previously shown that these MPs are suitable to stimulate T cell responses from either COVID-19 exposed or SARS-CoV-2-unexposed individuals (*27, 30*).

*Cytomegalovirus (CMV) MP*

As a control we have utilized a MP of 313 experimentally defined epitopes. This CMV MP consists of HLA Class-I and Class-II epitopes and $CD4^+$ and $CD8^+$ T cells have the capacity to recognize this MP, as have been previously published (*48*).

Flow cytometry assays

*Activation induced markers (AIM) assay*

Antigen-specific $CD4^+$ T cells were measured as a percentage of $AIM^+$ ($OX40^+CD137^+$) $CD4^+$ and ($CD69^+CD137^+$) $CD8^+$ T cells after stimulation of PBMCs from mRNA-1273 vaccinees and COVID-19 convalescent donors with peptide MPs. Also, antigen-specific circulating T follicular helper ($cT_{FH}$) cells ($CXCR5^+OX40^+CD40L^+$, as % of $CD4^+$ T cells) were defined by the AIM assay.

Prior to addition of MPs, cells were blocked at 37°C for 15 minutes with 0.5 µg/mL anti-CD40 mAb (Miltenyi Biotec). Then, cells were incubated at 37°C for 24 hours in the presence of fluorescently-labeled chemokine receptor antibodies (CCR6, CXCR5, CXCR3, CCR7, and CCR4) and SARS-CoV-2 MPs [1 µg/mL] or CMV MP [1 µg/mL] in 96-wells U bottom plates, as previously described (*27, 44*). In addition, PBMCs were incubated with an equimolar amount of DMSO as negative control and with phytohemagglutinin [5 µg/mL] (PHA, Roche) as a positive control.

For the surface stain, $1x10^6$ PBMCs were resuspended in PBS, incubated with BD human FC block (BD Biosciences, San Diego, CA) and the LIVE/DEAD marker in the dark for 15 min and washed with PBS. Then, antibody mix containing the rest of the surface antibodies were added directly to cells and incubated for 60 min at 4°C in the dark. Following surface staining, cells were then washed twice with PBS containing 3% FBS (FACS buffer). All samples were acquired on a Cytek Aurora (Cytek Biosciences, Fremont, CA). A list of antibodies used in this panel can be found in **Table S3** and a representative gating strategy of Spike-specific $CD4^+$ and $CD8^+$ T cells using the AIM assay is shown in **Figure S2**.

Antigen-specific $CD4^+$ and $CD8^+$ T cells were measured as background (DMSO) subtracted data, with a minimal DMSO level set to 0.005%. Response > 0.02% and a stimulation index (SI) > 2 for $CD4^+$ and > 0.03% and SI > 3 for $CD8^+$ T cells were considered positive. The limit of quantification (LOQ) for antigen-specific $CD4^+$ T cell responses (0.03%) and antigen-specific $CD8^+$ T cell responses (0.06%) was calculated using the median two-fold standard deviation of all negative controls. As internal quality control to define inter-assay variation, the CMV-specific $CD4^+$ and $CD8^+$ T cell responses were evaluated for a SARS-CoV-2-unexposed donor included in each independent experiments. The antigen-specific response against CMV and the response to the positive control was compared across experiments and revealed a coefficient of variation (CV) between 10 to 13% for the antigen-specific stimulation with the CMV-MP, and a CV between 6 to 12% for mitogenic stimulation with PHA **(Fig. S2)**.

*Intracellular cytokine staining (ICS) assay*

To optimize Spike-specific detection of cytokine-producing $CD4^+$ and $CD8^+$ T cells, we experimented with different incubation times (5, 6, and 24 hours) with the addition of Golgi-Plug containing brefeldin A and Golgi-Stop containing monensin (BD Biosciences, San Diego, CA) for an additional 1 or 3 hours at the beginning of the incubation time, or 4 hours at the end of the incubation time, respectively. To establish optimal conditions for the ICS assay, we evaluated the IFNγ-producing $CD4^+$ and $CD8^+$ T cells in 100 μg mRNA-1273 vaccinees (n=4) and COVID-19 convalescent donors (n=4) (**Table S5**). The highest signal of IFNγ-producing $CD4^+$ and $CD8^+$ T cells was detected after 24+4 hours incubation in both, vaccinees and convalescent donors. Hence, we chose 24 hours as the best condition to identify Spike-specific $CD4^+$ and $CD8^+$ T cells producing intracellular cytokines.

Prior to addition of MPs, cells were blocked at 37°C for 15 minutes with 0.5 μg/mL anti-CD40 mAb, as previously described (*44*). PBMCs were cultured in the presence of SARS-CoV-2 MPs [1 μg/mL] for 24 hours at 37°C. In addition, PBMCs were incubated with an equimolar amount of DMSO as negative control and also CMV MP [1 μg/mL] as a positive control. After the 24 hours, Golgi-Plug and Golgi-Stop were added to the culture for 4 hours, as described above. Cells were then washed and surface stained for 30 min at 4°C in the dark and fixed with 1% of paraformaldehyde (Sigma-Aldrich, St. Louis, MO). Antibodies used in the ICS assay are listed in **Table S4** and a representative gating strategy of Spike-specific $CD4^+$ and $CD8^+$ T cells using the ICS assay is shown in **Figure S4**.

Antigen-specific $CD4^+$ and $CD8^+$ T cells were measured as background (DMSO) subtracted data, with a minimal DMSO level set to 0.001%. Responses > 0.005% and a SI > 2 for $CD4^+$ and $CD8^+$ T cells was considered positive. The limit of quantification for antigen-specific $CD4^+$ and $CD8^+$ T cell responses (0.01%) was calculated using the median two-fold standard deviation of all negative controls. As internal quality control to define inter-assay variation, the CMV-specific $CD4^+$ and $CD8^+$ T cell responses were evaluated for a SARS-CoV-2-unexposed donor included in each independent experiment. The antigen-specific response of $CD4^+$ and $CD8^+$ T cells against

CMV were compared across experiments and revealed a CV of 14% for the antigen-specific stimulation with the CMV MP **(Fig. S4)**.

The gates applied for the identification of $CD4^+$ and $CD8^+$ T cells producing cytokines were defined according to the cells cultured with DMSO for each individual as is shown in Fig. S4. A Boolean analysis was carried out to define the multifunctional profiles on FlowJo 10.7.1 and the analysis included CD40L, GzB, IFNγ, IL-2, and TNFα gated on $CD3^+CD4^+$ cells and GzB, IFNγ, IL-2, and TNFα gated on $CD3^+CD8^+$ cells. To define the multifunctional profiles of antigen-specific T cells, all positive background-subtracted data (> 0.005% and a SI > 2 for $CD4^+$ T cells and > 0.002% and a SI > 2 for $CD8^+$ T cells) was aggregated into a combined sum of antigen-specific $CD4^+$ or $CD8^+$ T cells based on the number of functions. Values higher than the LOQ (0.01%) were considered for the analysis of the multifunctional antigen-specific T cell responses. The average of the relative $CD4^+$ and $CD8^+$ T cell response was calculated per group to define the proportion of multifunctional antigen-specific T cell responses (**Fig. S6** and **S8**).

Statistical analysis

Data and statistical analyses were done in FlowJo 10.7.1 and GraphPad Prism 8.4, unless otherwise stated. The statistical details of the experiments are provided in the respective figure legends. Data plotted in linear scale were expressed as Mean ± Standard Deviation (SD). Data plotted in logarithmic scales were expressed as Geometric Mean ± Geometric Standard Deviation (SD). Mann-Whitney or Wilcoxon tests were applied for unpaired or paired comparisons, respectively. Details pertaining to significance are also noted in the respective legends.

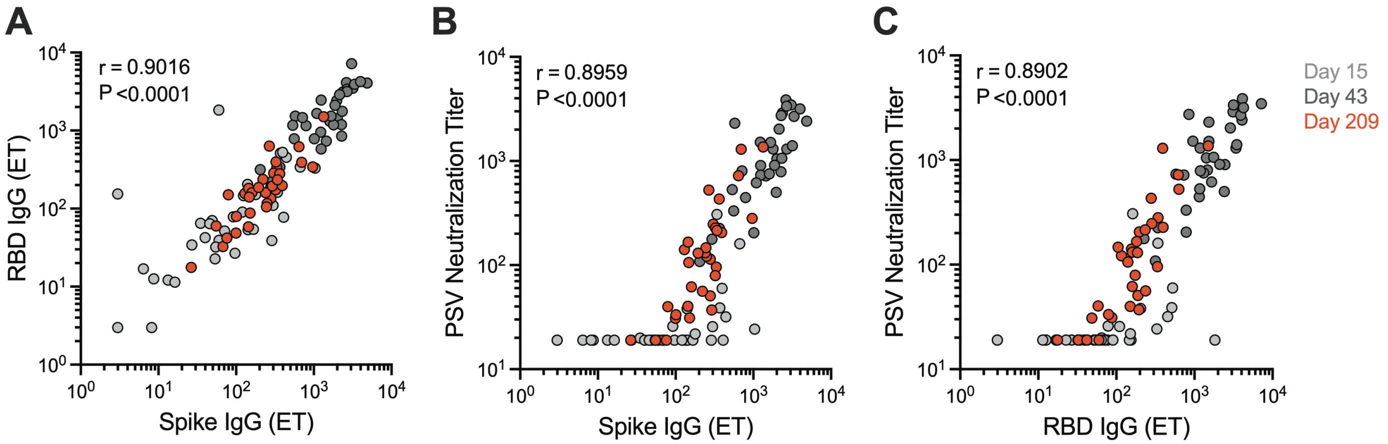


**Fig. S1. Correlation of antibody responses in mRNA-1273 vaccinees.**
**(A)** Correlation of Spike IgG and RBD IgG titers in mRNA-1273 vaccinees on days 15 ± 2, 43 ± 2, and 209 ± 7. **(B)** Correlation of Spike IgG and PSV neutralization titers in mRNA-1273 vaccinees on days 15 ± 2, 43 ± 2, and 209 ± 7. **(C)** Correlation of RBD IgG and PSV neutralization titers in mRNA-1273 vaccinees on days 15 ± 2, 43 ± 2, and 209 ± 7. Background-subtracted and log data analyzed in all cases. Day 15 ± 2 = light gray, day 43 ± 2 = dark gray, day 209 ± 7 = red. Statistics by Spearman's Rank Correlation test.

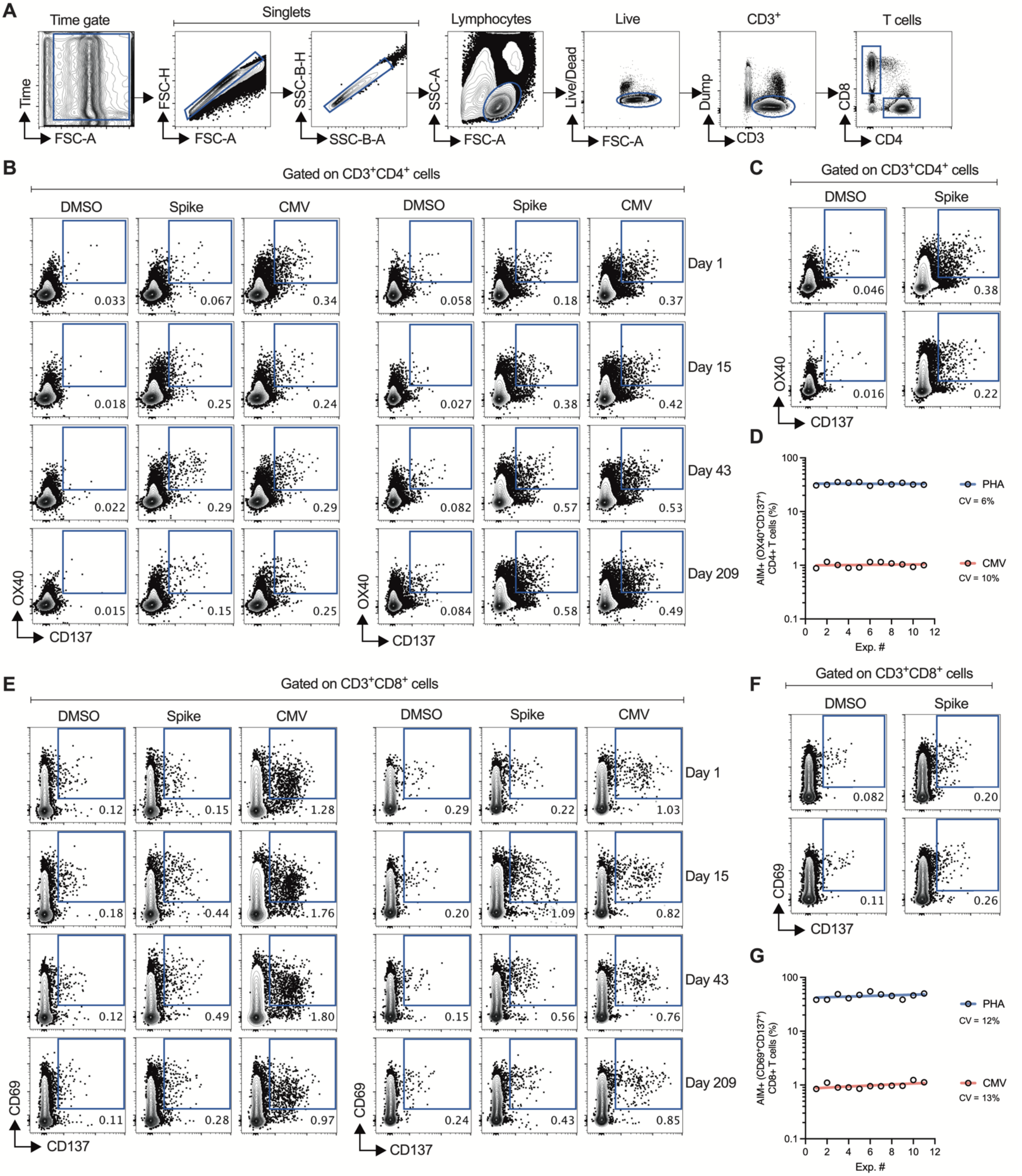

A
Time gate
Singlets
Lymphocytes
Live
CD3+
T cells
Time
FSC-A
FSC-H
FSC-A
SSC-B-H
SSC-B-A
SSC-A
FSC-A
Live/Dead
FSC-A
Dump
CD3
CD8
CD4
B
Gated on CD3+CD4+ cells
DMSO
Spike
CMV
DMSO
Spike
CMV
Day 1
0.033
0.067
0.34
0.058
0.18
0.37
Day 15
0.018
0.25
0.24
0.027
0.38
0.42
Day 43
0.022
0.29
0.29
0.082
0.57
0.53
Day 209
0.015
0.15
0.25
0.084
0.58
0.49
OX40
CD137
OX40
CD137
C
Gated on CD3+CD4+ cells
DMSO
Spike
0.046
0.38
0.016
0.22
OX40
CD137
D
AIM+ (OX40+CD137+) CD4+ T cells (%)
100
10
1
0.1
0
2
4
6
8
10
12
Exp. #
PHA
CV = 6%
CMV
CV = 10%
E
Gated on CD3+CD8+ cells
DMSO
Spike
CMV
DMSO
Spike
CMV
Day 1
0.12
0.15
1.28
0.29
0.22
1.03
Day 15
0.18
0.44
1.76
0.20
1.09
0.82
Day 43
0.12
0.49
1.80
0.15
0.56
0.76
Day 209
0.11
0.28
0.97
0.24
0.43
0.85
CD69
CD137
CD69
CD137
F
Gated on CD3+CD8+ cells
DMSO
Spike
0.082
0.20
0.11
0.26
CD69
CD137
G
AIM+ (CD69+CD137+) CD8+ T cells (%)
100
10
1
0.1
0
2
4
6
8
10
12
Exp. #
PHA
CV = 12%
CMV
CV = 13%

**Fig. S2. Assessment of Spike-specific T cells using an Activation Induced Marker (AIM) assay.**

**(A)** Gating strategies to define $CD3^+CD4^+$ and $CD3^+CD8^+$ cells by AIM assay. **(B and C)** Representative examples of flow cytometry plots of Spike-specific $CD4^+$ T cells ($OX40^+CD137^+$, after overnight stimulation with Spike megapool (MP) or CMV MP), compared to negative control (DMSO). FACS examples from two mRNA-1273 vaccinees (B) and two COVID-19 convalescent subjects (C). **(D)** CMV-specific $CD4^+$ T cell responses evaluated for a SARS-CoV-2-unexposed donor in the AIM experiments. This sample was included as internal quality control to define inter-assay variation. The antigen-specific response against CMV (red line) and the response to the positive control (PHA; blue line) was compared across the eleven independent experiments and revealed a coefficient of variation (CV) of 6% for mitogenic stimulation with PHA and a CV of 10% for the antigen-specific stimulation with the CMV MP. **(E and F)** Representative examples of flow cytometry plots of Spike-specific $CD8^+$ T cells ($CD69^+CD137^+$, after overnight stimulation with Spike MP or CMV MP), compared to DMSO. FACS examples from two mRNA-1273 vaccinees (E) and two COVID-19 convalescent subjects (F). **(G)** CMV-specific $CD8^+$ T cell responses evaluated for a SARS-CoV-2-unexposed donor in the AIM experiments. This sample was included as internal quality control to define inter-assay variation. Similar to the strategy used for the $AIM^+$ $CD4^+$ T cells, the antigen-specific response against CMV (red line) and the response to the positive control (PHA; blue line) was compared across the eleven independent experiments and revealed a CV of 12% for mitogenic stimulation with PHA and a CV of 13% for the antigen-specific stimulation with the CMV MP. Background-subtracted and log data analyzed in D and G.

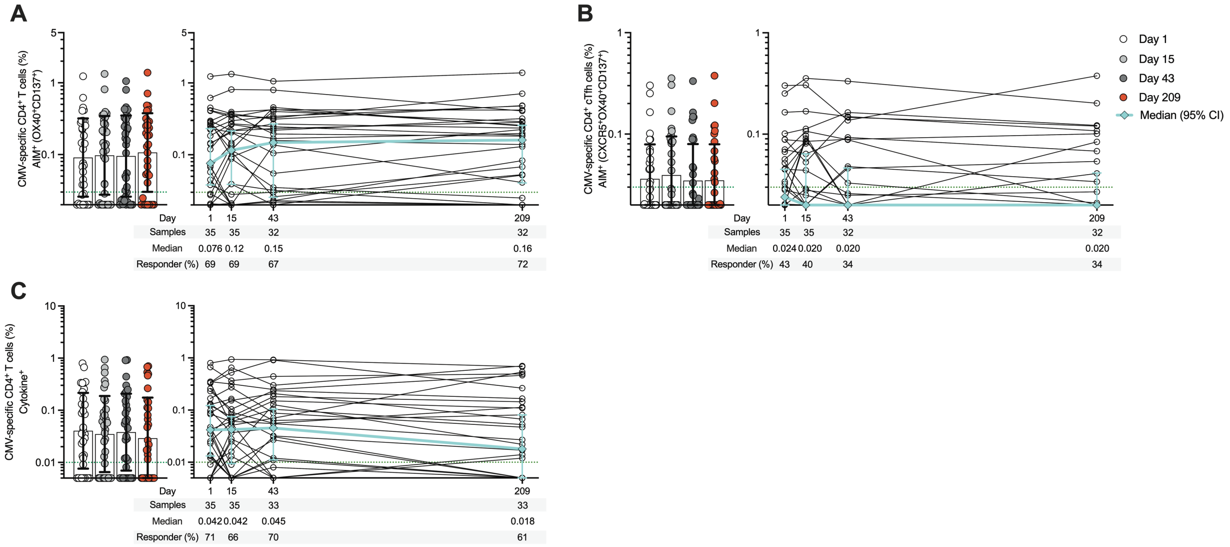


**Fig. S3. CMV-specific CD4$^+$ T cell responses in mRNA-1273 vaccinees.**

**(A)** Longitudinal CMV-specific CD4$^+$ AIM$^+$ T cells in mRNA-1273 vaccinees. Percentage of background subtracted CMV-specific CD4$^+$ T cells quantified by AIM (OX40$^+$CD137$^+$) after stimulation with CMV MP in mRNA-1273 vaccinees. **(B)** Quantitation of CMV-specific circulating T follicular helper (cT$_{FH}$) cells (CXCR5$^+$OX40$^+$sCD40L$^+$, as % of CD4$^+$ T cells) after overnight stimulation with CMV MP. **(C)** Longitudinal CMV-specific CD4$^+$ Cytokine$^+$ T cells expressing intracellular CD40L or producing IFNγ, TNFα, IL-2 or GzB in mRNA-1273 vaccinees. Percentage of background subtracted CMV-specific CD4$^+$ T cells quantified by ICS after stimulation with the CMV MP in mRNA-1273 vaccinees. A Boolean gating strategy was used to define the frequencies of CD4$^+$ T cells producing either IFNγ, TNFα, IL-2 or GzB or expressing CD40L. The dotted green line indicates limit of quantification (LOQ). The bars in A, B, and C indicate the geometric mean in the analysis of the CMV-specific CD4$^+$ T cell frequencies on days 1, 15 ± 2, 43 ± 2, and 209 ± 7 postimmunization. Background-subtracted and log data analyzed in all cases. The bottom panels in A, B, and C show the samples included in each group, the median of the frequencies, and the percentage of responders. Day 1 = white, day 15 ± 2 = light gray, day 43 ± 2 = dark gray, day 209 ± 7 = red. Statistics by Wilcoxon signed-rank test.

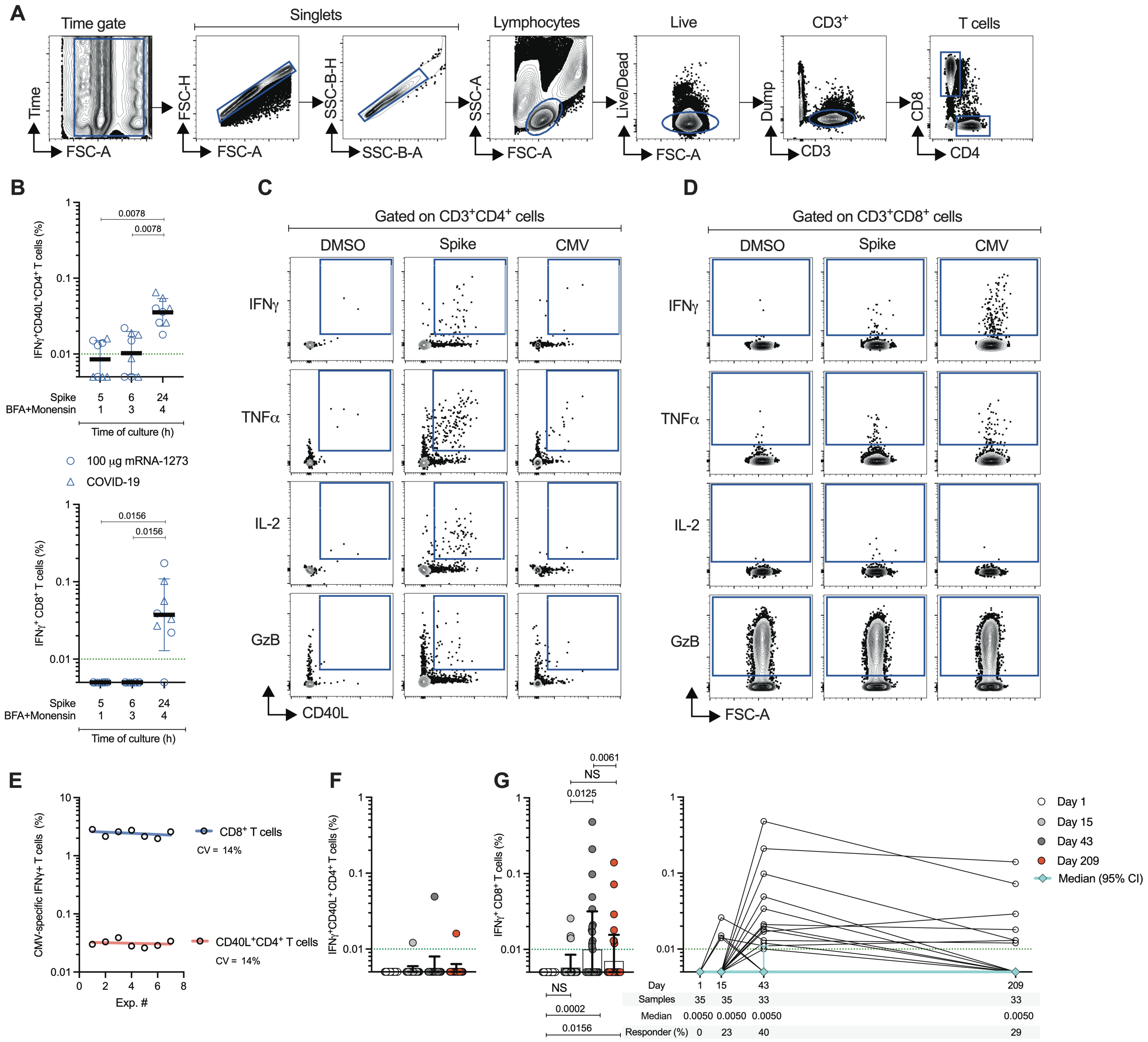


**Fig. S4. Assessment of Spike-specific T cells using an Intracellular Cytokine (ICS) assay.** **(A)** Gating strategies to define $CD3^+CD4^+$ and $CD3^+CD8^+$ cells by the ICS assay. **(B)** Spike-specific $CD4^+$ and $CD8^+$ T cells producing IFNγ at 5, 6, and 24 hours of stimulation with Spike MP. To optimize Spike-specific detection of cytokine-producing $CD4^+$ and $CD8^+$ T cells, we experimented with different incubation times (5, 6, and 24 hours) with the addition of brefeldin A (BFA) and monensin for an additional 1 or 3 at the beginning of the incubation time, or 4 hours at the end of the incubation time, respectively. We evaluated the IFNγ-producing $CD4^+$ and $CD8^+$ T cells in 100 µg mRNA-1273 vaccinees (n=4) and COVID-19 convalescent donors (n=4) (Table S5). The highest signal of IFNγ-producing T cells was detected after 24+4 hours for both Spike-specific $CD4^+$ and $CD8^+$ T cells (upper and lower panel, respectively) in vaccinees and convalescent donors. Hence, we chose 24 hours as the best condition to identify Spike-specific

$CD4^+$ and $CD8^+$ T cells producing cytokines. **(C and D)** Representative examples of flow cytometry plots of Spike-specific $CD4^+CD40L^+$ or $CD8^+$ T cells producing IFNγ, TNFα, IL-2, and GzB after overnight stimulation with Spike MP or CMV MP, compared to the negative control (DMSO). FACS examples from one mRNA-1273 vaccinees at day 43 ± 2. **(E)** CMV-specific $CD4^+CD40L^+$ and $CD8^+$ T cell responses evaluated for a SARS-CoV-2-unexposed donor in the AIM experiments. This sample was included as internal quality control to define the variation inter-assay. The CMV-specific $CD40L^+CD4^+$ and $CD8^+$ T cell response measured by IFNγ-production was compared across the seven independent experiments and revealed a coefficient of variation (CV) of 14% for $CD4^+$ and $CD8^+$ T cells. **(F and G)** Spike-specific $CD8^+$ T cell responses with an optimal pool of Class-I epitopes (CD8 Spike MP) in mRNA-1273 vaccinees. To confirm that the Spike-specific $CD8^+$ T cell response observed in the mRNA-1273 vaccinees is also induced in the absence of the Spike-specific $CD4^+$ T cell response, we incubated PBMCs with the CD8 Spike MP to evaluate the IFNγ-producing T cells, as described above. **(F)** As excepted, CD8 Spike MP does not induce $CD4^+$ T cell response at any timepoint postimmunization in the 25 μg mRNA-1273 vaccinees. Only in 1/35 (0.35%) donor was detected the $CD4^+$ T cell response using the CD8 Spike MP (CD8-S MP), consisting of 9 and 10-mers, Class-I restricted epitopes. **(G)** Spike-specific $CD8^+$ T cell response using the optimal MP was detectable in 23% (8/35) of the subjects after the first immunization. The response was significantly increased after the second immunization at day 43 and day 209 and the Spike-specific $CD8^+$ T cell response was detected in 40% (14/35) and 29% (10/35) of the subjects, respectively. This response follows similar kinetics of what had been seen in Spike-specific $CD8^+$ T cells producing IFNγ in response to the overlapping Spike MP consisting of overlapping 15-mers (Fig. 3). Background-subtracted and log data analyzed in B, E, F, and G. The dotted green line indicates limit of quantification (LOQ). The bars in F and G indicate the geometric mean in the analysis of the Spike-specific $CD4^+$ and $CD8^+$ T cell frequencies on days 1, 15 ± 2, 43 ± 2, and 209 ± 7 postimmunization. The bottom panel in G shows the samples included in each group, the median of the frequencies, and the percentage of responders. Day 1 = white, day 15 ± 2 = light gray, day 43 ± 2 = dark gray, day 209 ± 7 = red. Statistics by (B) Mann-Whitney U test and (G) Wilcoxon signed-rank test.

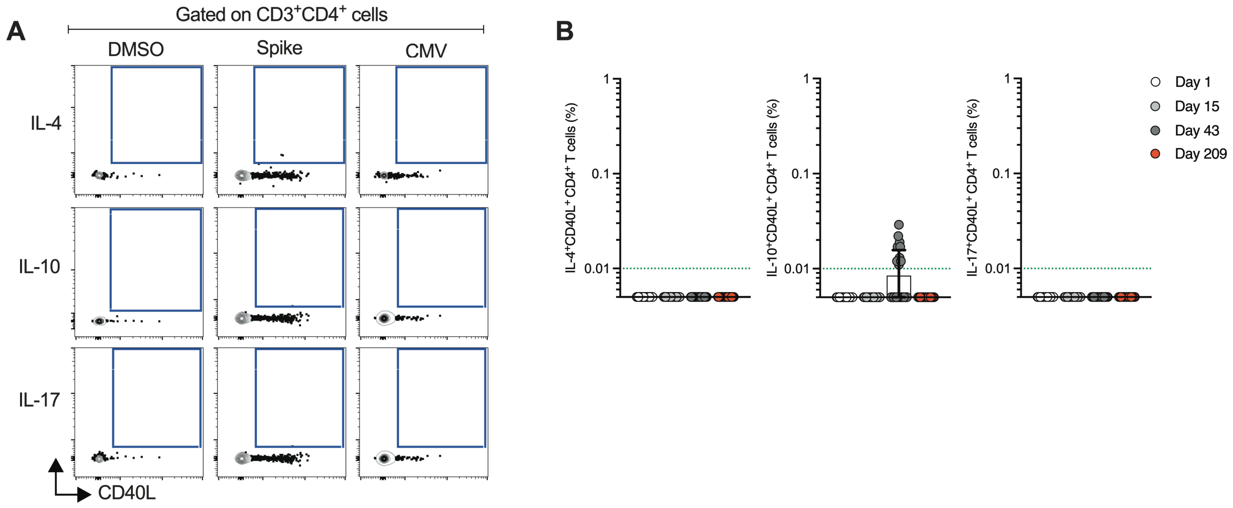


**Fig. S5. Spike-specific CD4$^+$ T cells producing IL-4, IL-10, and IL-17 in mRNA-1273 vaccinees.**

**(A)** Representative examples of flow cytometry plots of Spike-specific CD4$^+$CD40L$^+$ producing IL-4, IL-10, and IL-17 after overnight stimulation with Spike MP or CMV MP, compared to the negative control (DMSO). FACS examples from one mRNA-1273 vaccinees at day 43 ± 2. **(B)** Longitudinal Spike-specific CD4$^+$CD40L$^+$ T cells producing IL-4, IL-10, and IL-17 in mRNA-1273 vaccinees. No IL-4- or IL-17-producing CD4$^+$ T cells in response to Spike MP were detected at any timepoint postimmunization in 25 µg mRNA-1273 vaccinees. Spike-specific IL-10-producing CD4$^+$ T cell response was detected in 36% (12/33) of the subjects on day 43 while being undetectable in other timepoints. Day 1 = white, day 15 ± 2 = light gray, day 43 ± 2 = dark gray, day 209 ± 7 = red. Background-subtracted and log data analyzed in all cases. The dotted green line indicates limit of quantification (LOQ).

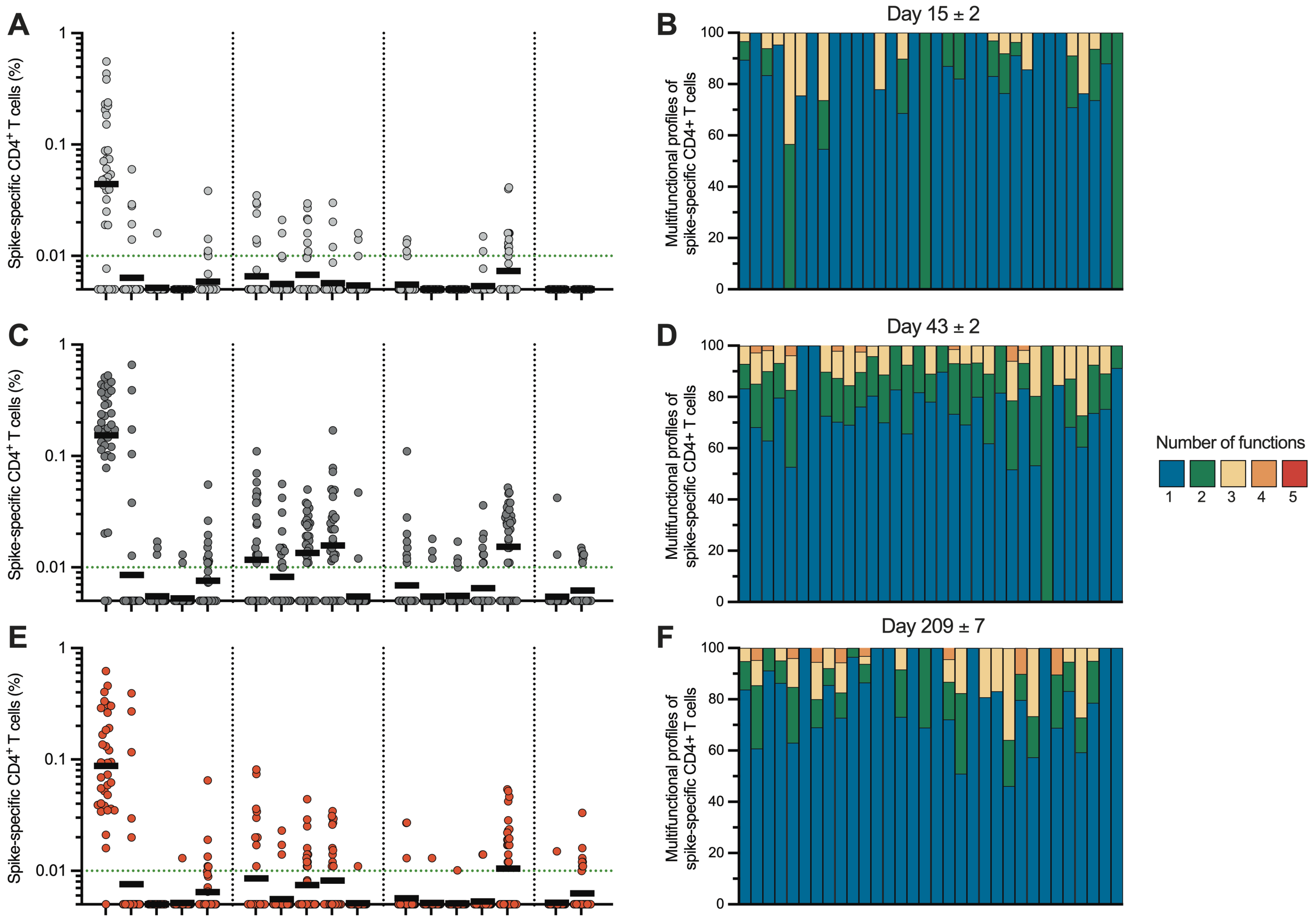


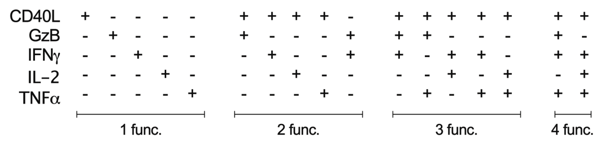


**G**

| | Day 15 ± 2 | | Day 43 ± 2 | | Day 209 ± 7 | |
|---|---|---|---|---|---|---|
| | Mean | SD | Mean | SD | Mean | SD |
| 1 Func. | 81.20 | 28.20 | 71.10 | 21.60 | 81.60 | 16.10 |
| 2 Func. | 12.70 | 25.00 | 17.70 | 16.60 | 10.00 | 9.59 |
| 3 Func. | 6.12 | 10.30 | 7.69 | 6.55 | 7.00 | 9.38 |
| 4 Func. | 0.00 | 0.00 | 3.48 | 16.80 | 1.39 | 2.85 |
| 5 Func. | 0.00 | 0.00 | 0.00 | 0.00 | 0.00 | 0.00 |

**Fig. S6. Multifunctional profiles of Spike-specific CD4$^+$ T cells in mRNA-1273 vaccinees. (A, C, E)** Predominant multifunctional profiles of Spike-specific CD4$^+$ T cells with one, two, three, and four functions were analyzed in mRNA-1273 vaccinees on days 15 ± 2 (A), 43 ± 2 (C), and 209 ± 7 (E) postimmunization. A Boolean analysis was carried out to define the multifunctional profiles on FlowJo 10.7.1 and the analysis included CD40L, GzB, IFNγ, IL-2, and TNFα gated on CD3$^+$CD4$^+$ cells (See Fig. S4). Background-subtracted and log data analyzed in all cases. The dotted green line indicates limit of quantification (LOQ). **(B, D, F)** Proportion of multifunctional Spike-specific CD4$^+$ T cells with one, two, three, and four functions in mRNA-1273 vaccinees on days 15 ± 2 (B), 43 ± 2 (D), and 209 ± 7 (F) postimmunization. Each bar shows the proportion of multifunctional Ag-specific CD4$^+$ T cells detected per donor. The blue, green, yellow, and orange colors in the pie charts depict the production of one, two, three, and four functions, respectively. **(G)** Mean and SD of multifunctional Spike-specific CD4$^+$ T cells with one, two, three, and four functions detected in mRNA-1273 vaccinees on days 15 ± 2, 43 ± 2, and 209 ± 7. Data plotted on Fig. 2F.

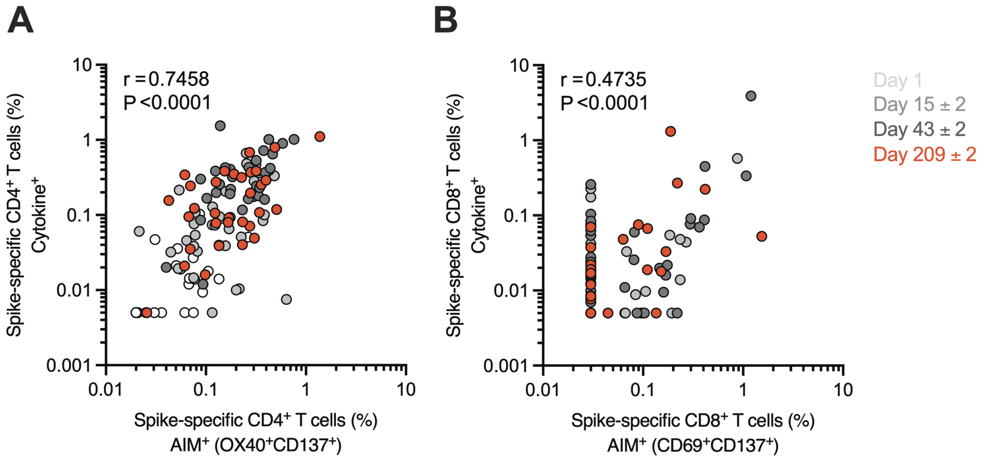


**Fig. S7. Correlation of Spike-specific CD4$^{+}$ and CD8$^{+}$ T cell response detected by AIM and ICS assays in mRNA-1273 vaccinees.**

**(A)** Correlation of Spike-specific CD4$^{+}$ T cell response using AIM and ICS assays in mRNA-1273 vaccinees. AIM$^{+}$CD4$^{+}$ T cell response was defined based on the double-expression of OX40$^{+}$CD137$^{+}$ (See Fig. S2) and ICS$^{+}$CD4$^{+}$ T cell response was defined based on the expression of CD40L or the production of IFNγ, TNFα, IL-2 or GzB (See Fig. S4). **(B)** Correlation of Spike-specific CD8$^{+}$ T cell response using AIM and ICS assays in mRNA-1273 vaccinees. AIM$^{+}$CD8$^{+}$ T cell response was defined based on the double-expression of CD69$^{+}$CD137$^{+}$ (See Fig. S2) and ICS$^{+}$CD8$^{+}$ T cell response was defined based on the production of IFNγ, TNFα, IL-2 or GzB (See Fig. S4). The response was analyzed on PBMCs after an overnight stimulation with overlapping Spike MP as described in the Materials and Methods section. Background-subtracted and log data analyzed in all cases. Day 1 = white, day 15 ± 2 = light gray, day 43 ± 2 = dark gray, day 209 ± 7 = red. Statistics by Spearman's Rank Correlation test.

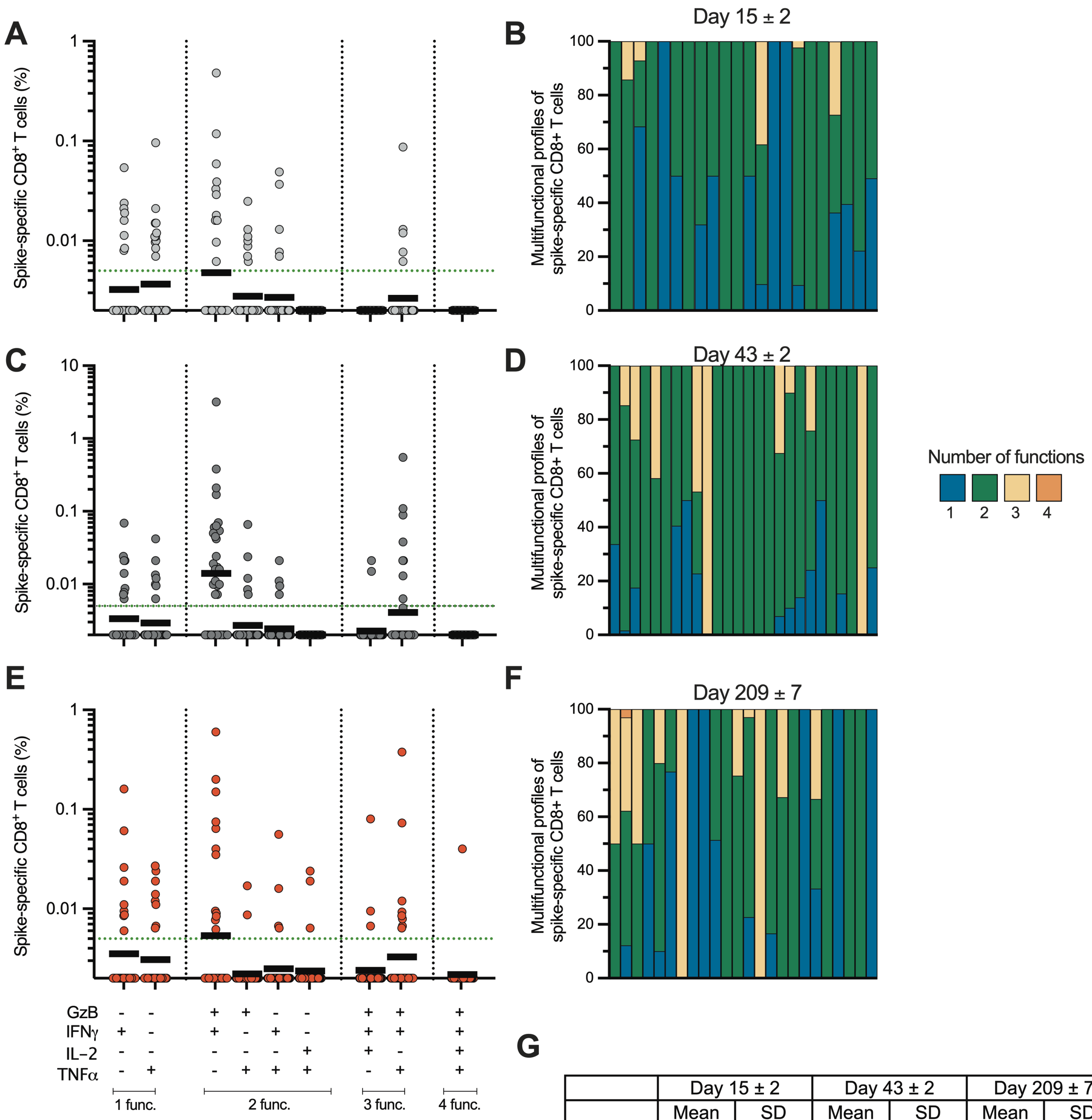


G

| | Day 15 ± 2 | | Day 43 ± 2 | | Day 209 ± 7 | |
|---|---|---|---|---|---|---|
| | Mean | SD | Mean | SD | Mean | SD |
| 1 Func. | 32.58 | 35.06 | 11.55 | 16.17 | 32.21 | 40.80 |
| 2 Func. | 63.36 | 35.09 | 73.73 | 29.99 | 48.97 | 38.46 |
| 3 Func. | 4.05 | 10.01 | 14.72 | 28.28 | 18.69 | 30.24 |
| 4 Func. | 0.00 | 0.00 | 0.00 | 0.00 | 0.13 | 0.62 |

**Fig. S8. Multifunctional profiles of Spike-specific CD8$^+$ T cells in mRNA-1273 vaccinees.** **(A, C, E)** Predominant multifunctional profiles of Spike-specific CD8$^+$ T cells with one, two, three, and four functions were analyzed in mRNA-1273 vaccinees on days 15 ± 2 (A), 43 ± 2 (C), and 209 ± 7 (E) postimmunization. A Boolean analysis was carried out to define the multifunctional profiles on FlowJo 10.7.1 and the analysis included GzB, IFNγ, IL-2, and TNFα gated on $CD3^+CD8^+$ cells (See Fig. S4). Background-subtracted and log data analyzed in all cases. The dotted green line indicates limit of quantification (LOQ). **(B, D, F)** Proportion of multifunctional Spike-specific $CD8^+$ T cells with one, two, three, and four functions in mRNA-1273 vaccinees on days 15 ± 2 (B), 43 ± 2 (D), and 209 ± 7 (F) postimmunization. Each bar shows the proportion of multifunctional Ag-specific $CD8^+$ T cells detected per donor. The blue, green, yellow, and orange colors in the pie charts depict the production of one, two, three, and four functions, respectively. **(G)** Mean and SD of multifunctional Spike-specific $CD8^+$ T cells with one, two, three, and four functions detected in mRNA-1273 vaccinees on days 15 ± 2, 43 ± 2, and 209 ± 7. Data plotted on Fig. 3E.

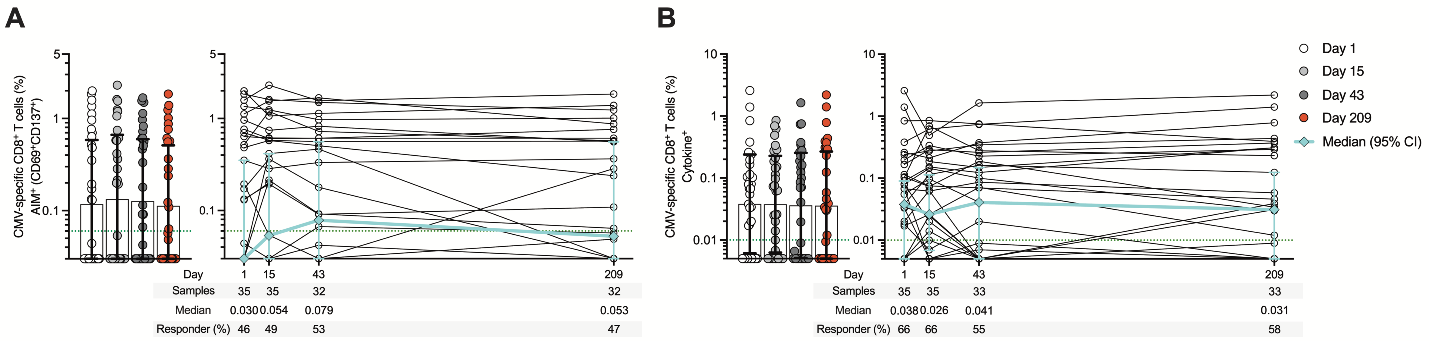


**Fig. S9. CMV-specific CD8$^+$ T cell responses in mRNA-1273 vaccinees.**
**(A)** Longitudinal CMV-specific CD8$^+$ AIM$^+$ T cells in mRNA-1273 vaccinees. Percentage of background subtracted CMV-specific CD8$^+$ T cells quantified by AIM (CD69$^+$CD137$^+$) after stimulation with CMV MP in mRNA-1273 vaccinees. **(B)** Longitudinal CMV-specific CD8$^+$ Cytokine$^+$ T cells producing IFNγ, TNFα, IL-2 or GzB in mRNA-1273 vaccinees. Percentage of background subtracted CMV-specific CD8$^+$ T cells quantified by ICS after stimulation with the CMV MP in mRNA-1273 vaccinees. A Boolean gating strategy was used to define the frequencies of CD8$^+$ T cells producing IFNγ, TNFα, IL-2 or GzB. The dotted green line indicates limit of quantification (LOQ). The bars in A and B indicate the geometric mean in the analysis of the CMV-specific CD8$^+$ T cell frequencies on days 1, 15 ± 2, 43 ± 2, and 209 ± 7 postimmunization. Background-subtracted and log data analyzed in all cases. The bottom panels in A and B show the samples included in each group, the median of the frequencies, and the percentage of responders. Day 1 = white, day 15 ± 2 = light gray, day 43 ± 2 = dark gray, day 209 ± 7 = red.

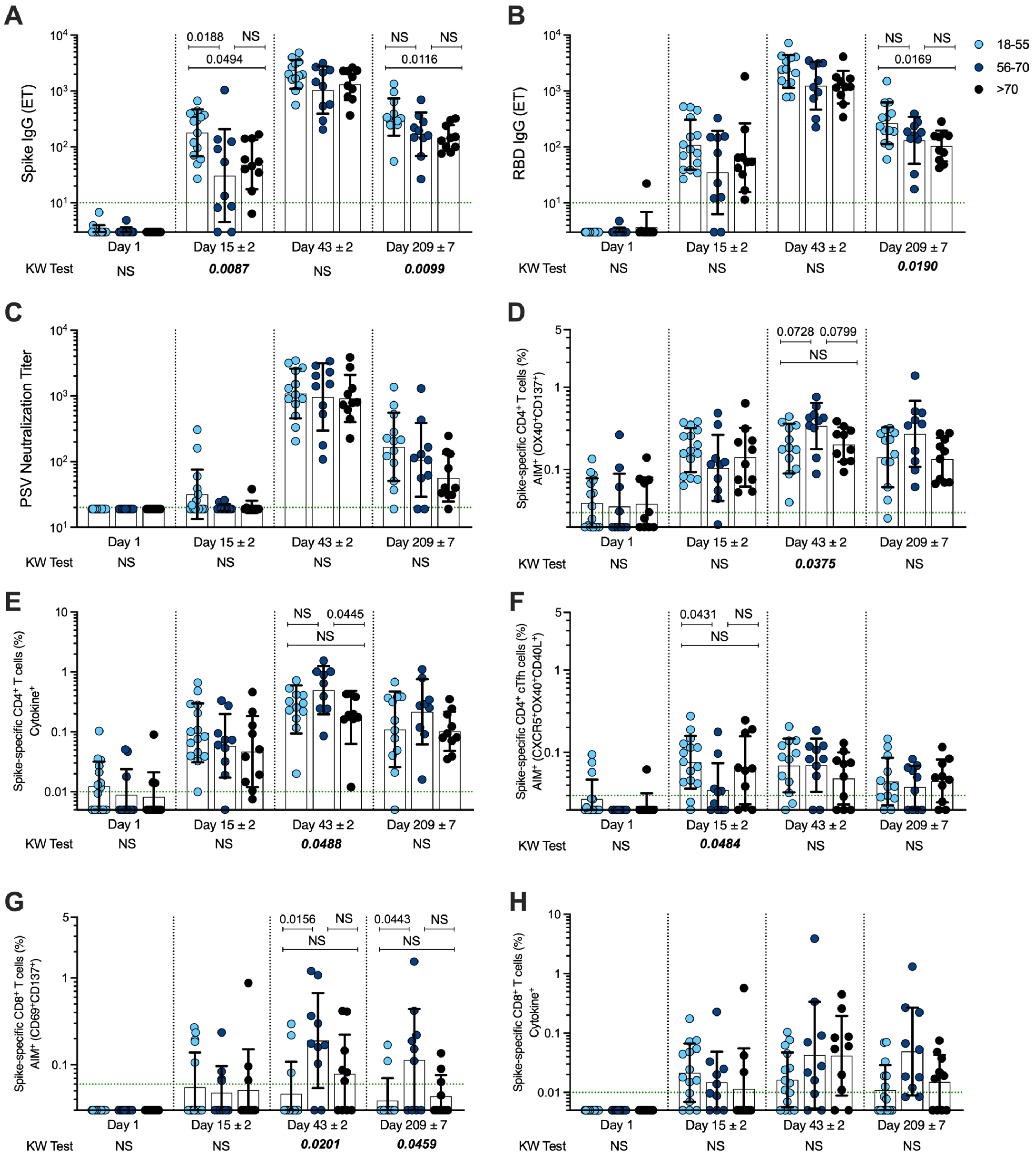

A
Spike IgG (ET)
0.0188
NS
0.0494
NS
NS
0.0116
Day 1
Day 15 ± 2
Day 43 ± 2
Day 209 ± 7
KW Test
NS
0.0087
NS
0.0099
B
RBD IgG (ET)
NS
NS
0.0169
18-55
56-70
>70
Day 1
Day 15 ± 2
Day 43 ± 2
Day 209 ± 7
KW Test
NS
NS
NS
0.0190
C
PSV Neutralization Titer
Day 1
Day 15 ± 2
Day 43 ± 2
Day 209 ± 7
KW Test
NS
NS
NS
NS
D
Spike-specific CD4+ T cells (%) AIM+ (OX40+CD137+)
0.0728
0.0799
NS
Day 1
Day 15 ± 2
Day 43 ± 2
Day 209 ± 7
KW Test
NS
NS
0.0375
NS
E
Spike-specific CD4+ T cells (%) Cytokine+
NS
0.0445
NS
Day 1
Day 15 ± 2
Day 43 ± 2
Day 209 ± 7
KW Test
NS
NS
0.0488
NS
F
Spike-specific CD4+ cTfh cells (%) AIM+ (CXCR5+OX40+CD40L+)
0.0431
NS
NS
Day 1
Day 15 ± 2
Day 43 ± 2
Day 209 ± 7
KW Test
NS
0.0484
NS
NS
G
Spike-specific CD8+ T cells (%) AIM+ (CD69+CD137+)
0.0156
NS
NS
0.0443
NS
NS
Day 1
Day 15 ± 2
Day 43 ± 2
Day 209 ± 7
KW Test
NS
NS
0.0201
0.0459
H
Spike-specific CD8+ T cells (%) Cytokine+
Day 1
Day 15 ± 2
Day 43 ± 2
Day 209 ± 7
KW Test
NS
NS
NS
NS

**Fig. S10. Spike-specific antibody and T cell responses are induced by mRNA-1273 vaccination in young and elderly individuals.**

**(A-C)** Spike IgG, RBD IgG, and PSV neutralizing titers in young, middle-aged, and elderly mRNA-1273 vaccinees. **(D-E)** Spike-specific $CD4^+$ T cells in young, middle-aged, and elderly mRNA-1273 vaccinees. Spike-specific $CD4^+$ T cells were analyzed by AIM (OX40+CD137+) (D) or ICS (producing either IFNγ, TNFα, IL-2, or GzB or expressing CD40L) assays (E). **(F)** Spike-specific circulating T follicular helper (cT$_{FH}$) cells ($CXCR5^+OX40^+CD40L^+$, as % of $CD4^+$ T cells) in young, middle-aged, and elderly mRNA-1273 vaccinees. **(G-H)** Spike-specific $CD8^+$ T cells in mRNA-1273 vaccinees evaluated in young, middle-aged, and elderly mRNA-1273 vaccinees. Spike-specific $CD8^+$ T cells were analyzed by AIM ($CD69^+CD137^+$) (G) or ICS (producing either IFNγ, TNFα, IL-2 or GzB) assays (H). Background-subtracted and log data analyzed in all cases. Young (18-55) = light blue, middle-aged (56-70) = dark blue, elderly individuals (>70) = black. Statistics by non-parametric ANOVA Kruskal–Wallis test and Dunn's posttest for multiple comparisons. The *p*-values plotted on the bottom show the Kruskal-Wallis test results, and the p values plotted on the top show the posttest analysis comparing age groups.

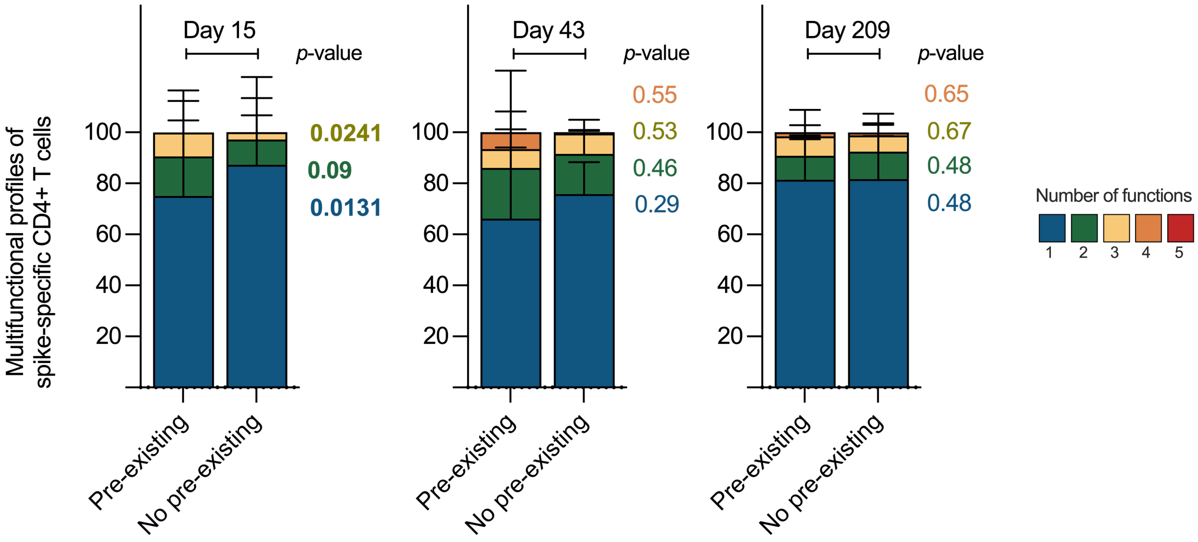


**Fig. S11. Multifunctional Spike-specific CD4$^+$ T cells in individuals with and without pre-existing spike immunity.**

Comparison of multifunctional profiles of Spike-specific CD4$^+$ T cells in individuals with and without pre-existing Spike immunity on days 15 ± 2, 43 ± 2, and 209 ± 7 postimmunization. The blue, green, yellow, and orange colors in the pie charts depict the production of one, two, three, and four functions, respectively. Statistics by Mann-Whitney U test. *p*-values coded by colors shown the comparison between individuals with and without pre-existing Spike immunity based on the number of functions of Spike-specific CD4$^+$ T cells.

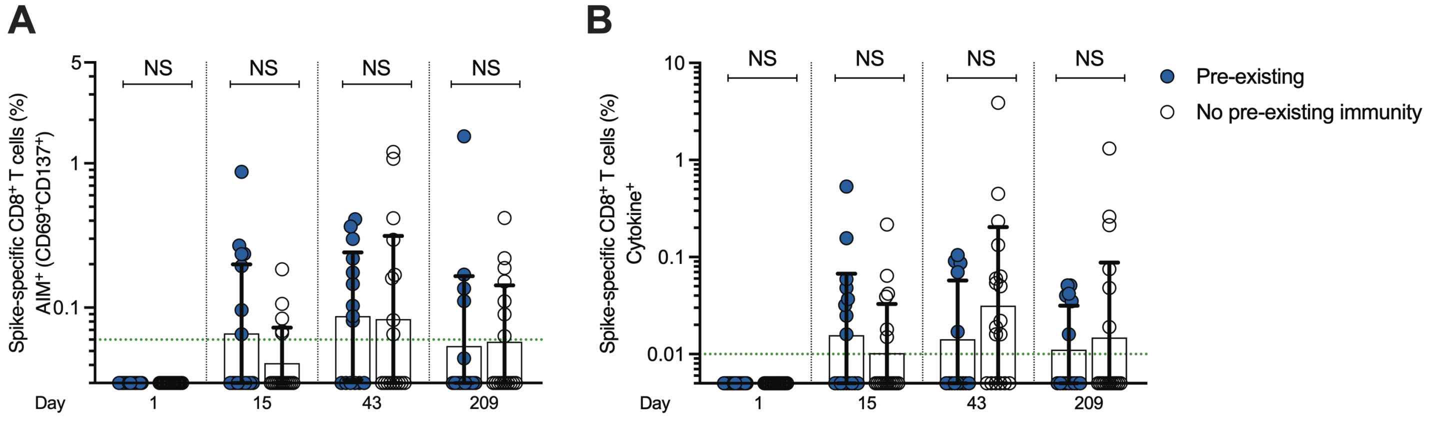


**Fig. S12. Spike-specific CD8$^+$ T cell response in not modulated by the pre-existing Spike immunity.**

**(A)** Spike-specific CD8$^+$ AIM$^+$ T cells in mRNA-1273 vaccinees with and without pre-existing Spike immunity evaluated on days 1, day 15 ± 2, day 43 ± 2, and day 209 ± 7 postimmunization. **(B)** Spike-specific CD8$^+$ Cytokine$^+$ T cells in mRNA-1273 vaccinees with and without pre-existing Spike immunity evaluated on days 1, day 15 ± 2, day 43 ± 2, and day 209 ± 7 postimmunization. The dotted green line indicates limit of quantification (LOQ). The bars in A and B indicate the geometric mean in the analysis of the Spike-specific CD8$^+$ T cells on day 1, 15 ± 2, 43 ± 2, and 209 ± 7 postimmunization. Background-subtracted and log data analyzed in all cases. Vaccinees with pre-existing Spike immunity = blue, vaccinees without pre-existing Spike immunity = clear. Statistics by Mann-Whitney U test. NS, non-significant.

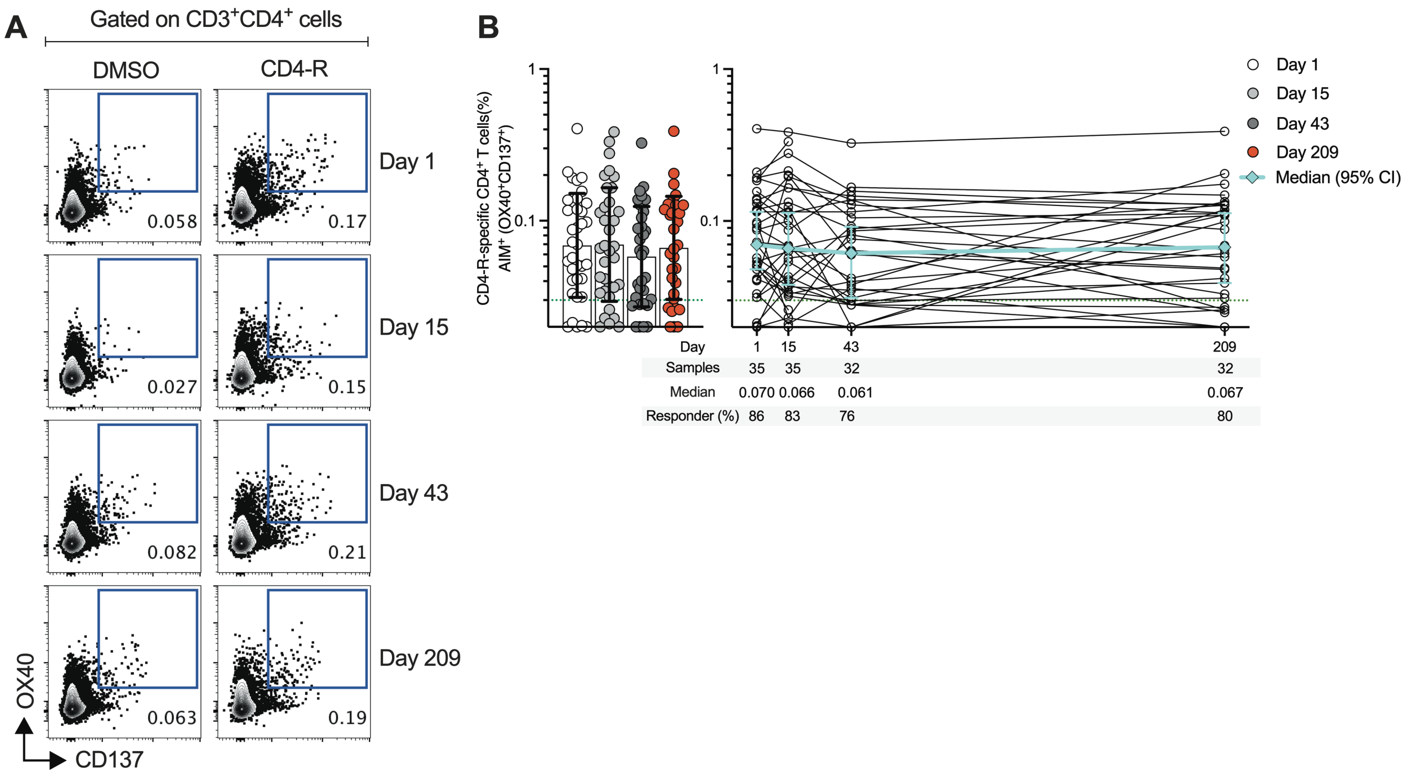


**Fig. S13. Non-Spike-specific CD4$^+$ T cells in mRNA-1273 vaccinees.**
**(A)** Representative examples of flow cytometry plots of CD4-R-specific CD4$^+$ T cells (OX40$^+$CD137$^+$, after overnight stimulation with CD4-R megapool (MP), compared to negative control (DMSO). FACS examples from one mRNA-1273 vaccinee. **(B)** Longitudinal CD4-R-specific CD4$^+$ AIM$^+$ T cells in mRNA-1273 vaccinees. The green line indicates limit of quantification (LOQ). The bar in B indicates the geometric mean in the analysis of the CD4-R-specific CD4$^+$ T cells on days 1, 15 ± 2, 43 ± 2, and 209 ± 7 postimmunization. Background-subtracted and log data analyzed in all cases. The bottom panel in B shows the samples included in each group, the median of the frequencies, and the percentage of responders. Day 1 = white, day 15 ± 2 = light gray, day 43 ± 2 = dark gray, day 209 ± 7 = red.

**Table S1. Characteristics of the 25 µg mRNA-1273 vaccinees.**

| Characteristic | 25 µg mRNA-1273 cohort | Age groups 18-55† | 56-70† | >70† |
|---|---|---|---|---|
| Donors, n | 35œ | 15 | 10 | 10 |
| Gender, n (%) | | | | |
| Male | 20 (57%) | 9 (60%) | 3 (30%) | 8 (80%) |
| Female | 15 (43%) | 6 (40%) | 7 (70%) | 2 (20%) |
| Age, years (mean ± SD) | 58.4 ± 19.1 | 36.7 ± 7.9 | 65.8 ± 4.5 | 72.8 ± 1.2 |
| Race or ethnicity, n (%)¥ | | | | |
| Asian | 1 | 0 | 0 | 1 (10) |
| White | 34 | 15 (100) | 10 (100) | 9 (90) |
| Hispanic or latino | 2 | 1 (7) | 0 | 1 (10) |

†Jackson, LA; et al., and Anderson, EJ; et al. reported the characteristics of the participants previously (Jackson and Anderson, NEJM)

¥Race or ethnic group could include more than one category per donor.

œTwo donors who did not receive the second dose were excluded from the analysis of the immune responses on days 43 and 209.

**Table S2. Characteristics of COVID-19 convalescent donors.**

| Characteristic | COVID-19 convalescent donors |
|---|---|
| Donors, n | 14 |
| Gender, n (%) | |
| Male | 4 (71%) |
| Female | 10 (29%) |
| Age, years (mean ± SD) | 34.5 ± 16.53 |
| Race or ethnicity, n (%)[¥] | |
| Asian | 0 |
| White | 13 (93%) |
| Hispanic or latino | 1 (7%) |
| SARS-CoV-2 PCR positivity, n (%) | |
| Positive | 14 (100%) |
| Negative | 0 |
| Days PSO at collection, Median (range) | 181 (170-195) |

PSO, post-symptom onset

**Table S3. Reagents used for AIM assays.**

| Reagent | Clone (Source) | Catalog. No. | Dilution |
|---|---|---|---|
| Live/Dead Blue | (ThermoFisher) | L23105 | 1:1000 |
| CCR6-BUV496 | 11A9 (BD Biosciences) | 612948 | 1:200 |
| CXCR5-BV421 | J252D4 (Biolegend) | 356920 | 1:200 |
| CXCR3-BV605 | G025H7 (Biolegend) | 353728 | 1:200 |
| CCR7-BV711 | G043H7 (Biolegend) | 353228 | 1:200 |
| CCR4-PE-Cy7 | L291H4 (Biolegend) | 359410 | 1:1000 |
| CD3-BUV395 | UCHT1 (BD Biosciences) | 563546 | 1:1000 |
| ICOS-BUV563 | DX29 (BD Biosciences) | 741421 | 1:200 |
| CD137-BUV737 | 4B4-1 (BD Biosciences) | 741861 | 1:100 |
| CD8-BUV805 | SK1 (BD Biosciences) | 612889 | 1:1000 |
| CD16-BV510 | 3G8 (Biolegend) | 302048 | 1:1000 |
| CD14-BV510 | 63D3 (Biolegend) | 367124 | 1:1000 |
| CD20-BV510 | 2H7 (Biolegend) | 302340 | 1:1000 |
| CD45RA-BV570 | HI100 (Biolegend) | 304132 | 1:1000 |
| CD38-BV650 | HB-7 (Biolegend) | 356620 | 1:200 |
| PD-1-BV785 | EH12.2H7 (Biolegend) | 329930 | 1:200 |
| CD69-FITC | FN50 (Biolegend) | 310904 | 1:200 |
| CD4-cFluor b548 | SK3 (Cytek Biosciences) | R7-20043 | 1:500 |
| CD95-BB700 | DX2 (BD Biosciences) | 566542 | 1:500 |
| PDL-1-PE | 29E.2A3 (Biolegend) | 329706 | 1:1000 |
| CD40L-PE-Dazzle594 | 24-31 (Biolegend) | 310840 | 1:200 |
| OX40-APC | Ber-Act35 (Biolegend) | 350008 | 1:100 |
| HLA-DR-APC-R700 | G46-6 (BD Biosciences) | 565127 | 1:500 |
| CD25-APC-Fire 750 | BC96 (Biolegend) | 302642 | 1:100 |

**Table S4. Reagents used for ICS assays.**

| Reagent | Clone (Source) | Catalog. No. | Dilution |
|---|---|---|---|
| Live/Dead Blue | (ThermoFisher) | L23105 | 1:1000 |
| CD3-BUV395 | UCHT1 (BD Biosciences) | 563546 | 1:100 |
| CD8-BUV805 | SK1 (BD Biosciences) | 612889 | 1:100 |
| CD16-BV510 | 3G8 (Biolegend) | 302048 | 1:200 |
| CD14-BV510 | 63D3 (Biolegend) | 367124 | 1:200 |
| CD20-BV510 | 2H7 (Biolegend) | 302340 | 1:200 |
| CD45RA-BV570 | HI100 (Biolegend) | 304132 | 1:50 |
| CD4-cFluor b548 | SK3 (Cytek Biosciences) | R7-20043 | 1:25 |
| CD95-PE-Cy5 | DX2 (BD Biosciences) | 559773 | 1:50 |
| CCR7-PE-Cy7 | G043H7 (Biolegend) | 353226 | 1:100 |
| IL-4-BUV737 | MP4-25D2 (BD Biosciences) | 612835 | 1:200 |
| IL-17-BV785 | BL168 (Biolegend) | 512338 | 1:100 |
| IFNγ-FITC | 4S.B3 (ThermoFisher) | 11-7319-82 | 1:500 |
| IL-2-BB700 | MQ1-17H12 (BD Biosciences) | 566405 | 1:200 |
| IL-10 -PE-Dazzle594 | JES3-19F1 (Biolegend) | 506812 | 1:100 |
| TNFα-eFluor450 | Mab11 (ThermoFisher) | 48-7349-42 | 1:200 |
| Granzyme B-AF647 | GB11 (BD Biosciences) | 560212 | 1:50 |
| CD40L-PerCP-ef710 | 24-31 (ThermoFisher) | 46-1548-42 | 1:50 |

**Table S5. Additional cohort of COVID-19 convalescent donors and 100 μg mRNA-1273 vaccinees included in the ICS assay[Δ].**

| Characteristic | COVID-19 convalescent donors | 100 μg mRNA-1273 vaccinees |
|---|---|---|
| Donors, n | 4 | 4 |
| Gender, n (%) | | |
| Male | 2 (50%) | 0 |
| Female | 2 (50%) | 4 (100%) |
| Age, year, median (range) | 51 (37-67) | 49.5 (41-59) |
| Race or ethnicity, n (%)[¥] | | |
| Asian | 0 | 2 (50%) |
| White | 3 (75%) | 2 (50%) |
| Hispanic or Latino | 0 | 0 |
| Unknown | 1 (25%) | 0 |
| SARS-CoV-2 PCR or Spike IgG positivity, n (%) | | |
| Positive | 4 (100%) | - |
| Negative | 0 | - |
| Days at collection, median (range) | | |
| Post-symptom onset | 91 (47-136) | - |
| Post-vaccination after first immunization[†] | - | 41 (40-47) |

[Δ]See Fig. S4 for additional information.

[†]Individuals received the second dose of 100 μg mRNA-1273 15 days before collection.
